# Canadian Association of Cardiovascular Prevention and Rehabilitation (CACPR) Exercise Training Recommendations in Cardiovascular Rehabilitation

**DOI:** 10.64898/2026.02.11.26346118

**Authors:** Diana Hopkins-Rosseel, Jennifer Harris, Paula Aver Bretanha Ribeiro, Simon L. Bacon, Nancy Hansen, Tim Hartley, Andree-Anne Hebert, Dustin E. Kimber, Billie-Jo Mabey, Ariany Marques Vieira, Stephanie Prince Ware, Patrick Warner, Kimberley Way, Colin Yeung

**Author notes:** ***Corresponding Author:*** Simon L Bacon, Concordia and CIUSSS-NIM (Hopital du Sacre-Coeur de Montreal) Concordia University: 7141 Sherbrooke St W, Montreal, Quebec H4B 1R6.

## Abstract

Exercise training is a cornerstone of Cardiovascular Rehabilitation (CR) and, as of now, moderate-to-vigorous continuous exercise training (MICT) is the standard. New exercise modalities in the context of CR are constantly being explored to improve patient outcomes. These Canadian Association of Cardiovascular Prevention and Rehabilitation (CACPR) exercise training recommendations provide a synthesis of evidence-informed recommendations from existing documents, including recommendations around High-Intensity Interval training (HIIT). CACPR created a pan-Canadian Exercise Working Group with various knowledge users (e.g., kinesiologists/exercise physiologists, physiotherapists, cardiologists, and patients) with expertise in CR-based exercise, who developed knowledge gap questions related to exercise training based on a literature review and synthesis of all available recommendations. An independent evidence-synthesis team performed a rapid review and meta-analyses to address the questions. The working group used this data to develop relevant recommendations. The final guidelines include 12 recommendations for CR exercise, including nine from previous documents and three new recommendations based on HIIT. The previous recommendations address exercise assessments and prescriptions for CR for various patient profiles. The new recommendations suggest that HIIT can be used to improve exercise capacity in patients with coronary artery disease (CAD), heart failure (HF) or atrial fibrillation. They also state that HIIT is superior to MICT in patients with CAD, that patients with HF should be considered for either HIIT or MICT and that any HIIT interval duration can be used as part of CR. Overall, these recommendations provide guidance for exercise in Canadian CR programs.

## 1. INTRODUCTION

Cardiovascular rehabilitation (CR) is a multi-component intervention that includes several risk-management strategies, such as exercise and activity training, psychosocial support, nutritional and vocational counseling, and patient education.^1^ CR has been shown to return patients to work, improve quality of life, positively impact healthcare costs and reduce cardiovascular mortality and morbidity.^2–5^ However, CR is still underutilised, with a global enrollment of 20-50%.^6–8^ Multiple CR-related associations and societies have published CR guidance documents. ^9–11^ Despite the abundance of guidance documents, very few describe the methods used to develop the recommendations, making it difficult to identify those that are based on high-quality evidence. In addition, very few guidance documents involve a diverse group of knowledge users in developing the guidelines.

The Canadian Association of Cardiovascular Prevention and Rehabilitation (CACPR) last released CR recommendations in 2009. This comprehensive document includes 165 formal recommendations across 16 chapters.^12^ Since then, there has been a notable expansion in CR research, necessitating the need to update these recommendations. After surveying CR experts and interested parties (e.g., patient partners, cardiologists) in Canada, exercise training was ranked as the first priority component to be updated. Exercise training is a cornerstone of CR,^1,13^ and various reviews have shown how exercise-based CR can positively impact patient outcomes, contributing to improvements in exercise capacity and decreases in hospitalisations and mortality.^2,13–15^ Most exercise recommendations include both continuous moderate-to-vigorous intensity aerobic exercise and resistance/flexibility exercises. More recently, there has been growing evidence around alternative exercise modalities, such as high-intensity interval training (HIIT), which were not addressed in the previous guidelines. Evidence suggests that HIIT may be more effective than MICT for improving peak oxygen uptake in patients with CAD, HF,^9,16^ or heart transplant.^17,18^ HIIT may also show greater benefits for left ventricular function and endothelial function in patients with HF.^19^

The goal of this updated CACPR exercise recommendations document was to provide the Canadian CR community with user-informed recommendations. To do this, existing evidence-informed recommendations were leveraged to consolidate prior recommendations. In addition, new recommendations that incorporated HIIT were created from a rapid review and meta-analyses. Collectively, this should provide Canadian CR programmes with high-quality guidance.

## 2. METHODS

### 2.1 Overview of the process

CACPR created a Guidelines Executive Committee, consisting of a Pan-Canadian group of experts, to oversee the guidelines and recommendations process. Their initial step was to survey the community to identify the priority areas for guideline updates, a process which identified exercise as the highest priority area. The executive committee then selected an Exercise Working Group to oversee the update using an open application process. Details of this process are available in the Supplement (Overview of the Project). To support the Exercise Working Group the executive committee engaged an evidence synthesis team (the Montreal Behavioural Medicine Centre’s META group) to undertake a review of existing guidance documents and conduct evidence syntheses on areas to be included in the updated recommendations.

### 2.2 The Exercise Working Group

The 11 members of the pan-Canadian Exercise Working Group included both patients and CR/exercise-related clinical experts from various disciplines (see Supplement, Table S1). This working group reviewed the evidence summary that was generated from previous recommendation documents (see the report of the SR of exercise recommendation https://osf.io/9ykrp), identifying evidence-informed recommendations. They also identified gaps in the recommendations and key questions that need to be addressed in the current clinical context to generate updated recommendations.

### 2.3 Evidence level grade coding

All recommendations that were generated (either from existing documents or newly generated) were coded using the Hypertension Canada Guidelines grading system:20

- **Grade A** - Recommendations are based on randomised trials (or systematic reviews of trials) with high levels of internal validity and statistical precision, and for which the study results can be directly applied to patients because of similar clinical characteristics and the clinical relevance of the study outcomes.
- **Grade B** - Recommendations are based on randomised trials, systematic reviews or pre-specified subgroup analyses of randomised trials that have lower precision, or there is a need to extrapolate from studies because of different populations or reporting of validated intermediate/surrogate outcomes rather than clinically important outcomes.
- **Grade C** - Recommendations are based on trials that have lower levels of internal validity and/or precision, trials reporting un-validated surrogate outcomes, or results from non-randomized observational studies.
- **Grade D** - Recommendations are on the basis of expert opinion alone or retrospective data only.

The questions identified by the working group that did not have evidence to support the creation of at least GRADE C recommendations, or areas not addressed by the evidence, were turned into recommendations for future research. Any recommendation that obtained a D grade was automatically excluded from this guidance document.

### 2.4 Systematic review of previous CR guidance documents

#### 2.4.1 Search

The systematic review was developed according to the Cochrane handbook^21^ and reported according to the Preferred Reporting Items for Systematic Reviews and Meta-Analyses (PRISMA)^22^ to capture previous recommendation documents focused on CR. An experienced health science librarian reviewed the search strategy. CINAHL, Embase, NICE, Ovid Medline, Pedro, Scielo, CPG Infobase, and Prospero were searched from 2004 through July 2019 with an update in August 2020 to identify evidence-based guidelines, position statements, and expert or clinical consensus documents (full details are available in the Guidelines Report; https://osf.io/9ykrp). An extensive search on the grey literature of clinical associations’ websites in the cardiovascular field was also undertaken. There were no search limits concerning language; however, due to the capacity of the evidence synthesis team, only documents written in English, French, Portuguese, Italian, and Spanish were considered for inclusion.

#### 2.4.2 Study selection

From the retrieves, we retained documents that presented evidence-informed exercise-related recommendations. Evidence-informed was defined as being developed based on a systematic review of the literature, with the authors grading the evidence to form the recommendations. Documents that provide recommendations on CR or secondary prevention for adults with a history of cardiovascular disease (e.g., guidelines, position statements and/or expert or clinical consensuses documents) were included. Cardiovascular disease was defined as: acute myocardial infarction; acute coronary syndrome; percutaneous revascularisation; coronary interventions; arrhythmias; atrial fibrillation; valve disorders; vascular cardiac disorders; or heart failure. Documents that included only: primary prevention populations; pharmacological treatments; and rehabilitation focused on patients post-stroke were excluded. For the cases of coexistence with any of the morbidities in our inclusion list, the study was included.

#### 2.4.3 Data extraction

Extracted data was entered into a Microsoft Office Excel™ spreadsheet designed for this study. Data of interest included descriptive characteristics of the papers: year of publication; journal; author; title; country; details of the exercise recommendations; the name of the organisation endorsing the recommendations; summarised objective of the guidance document; the inclusion (or not) of conflicts of interests for those generating the document; sponsorship/financing; if the document recommendations were based on a systematic literature search; and if they provide information or recommendations on additional CR components beyond exercise (e.g., nutrition, smoking cessation, stress management). We only extracted recommendations on exercise and focused on the target patient populations described in this document, the target outcome, the strength of the recommendations, and the level of the evidence. We classified the recommendations into 5 categories: patient assessment; aerobic exercise prescription/training; resistance exercise prescription/training; flexibility exercises; and patient safety.

### 2.5 Rapid Review for new recommendations

#### 2.5.1 Development of potential research questions

Based on the report of the available guidelines, along with the clinical experience and knowledge of the members, the Exercise Working Group generated a list of exercise prescription/training-related questions that needed to be addressed. The group identified HIIT as the most pressing area for further exploration. Traditionally, moderate-to-vigorous intensity continuous training (MICT) is prescribed for patients with cardiovascular disease^9,23^ but there is a recent growing evidence base to support the use of HIIT in CR. HIIT is a prescription modality for aerobic training that intersperses periods of relatively higher intensity exercise (e.g., ≥75% of maximal capacity) with recovery periods that can vary from very low intensity exercise to no activity/movement.^24,25^

The following clinical questions (Box 1) were developed by the working group in collaboration with the META group. The detailed description of the search, selection, and extraction process are described in an additional report (HIIT Report available at https://osf.io/73g4w/).

#### Box 1.

##### Clinical questions that guided the new recommendations

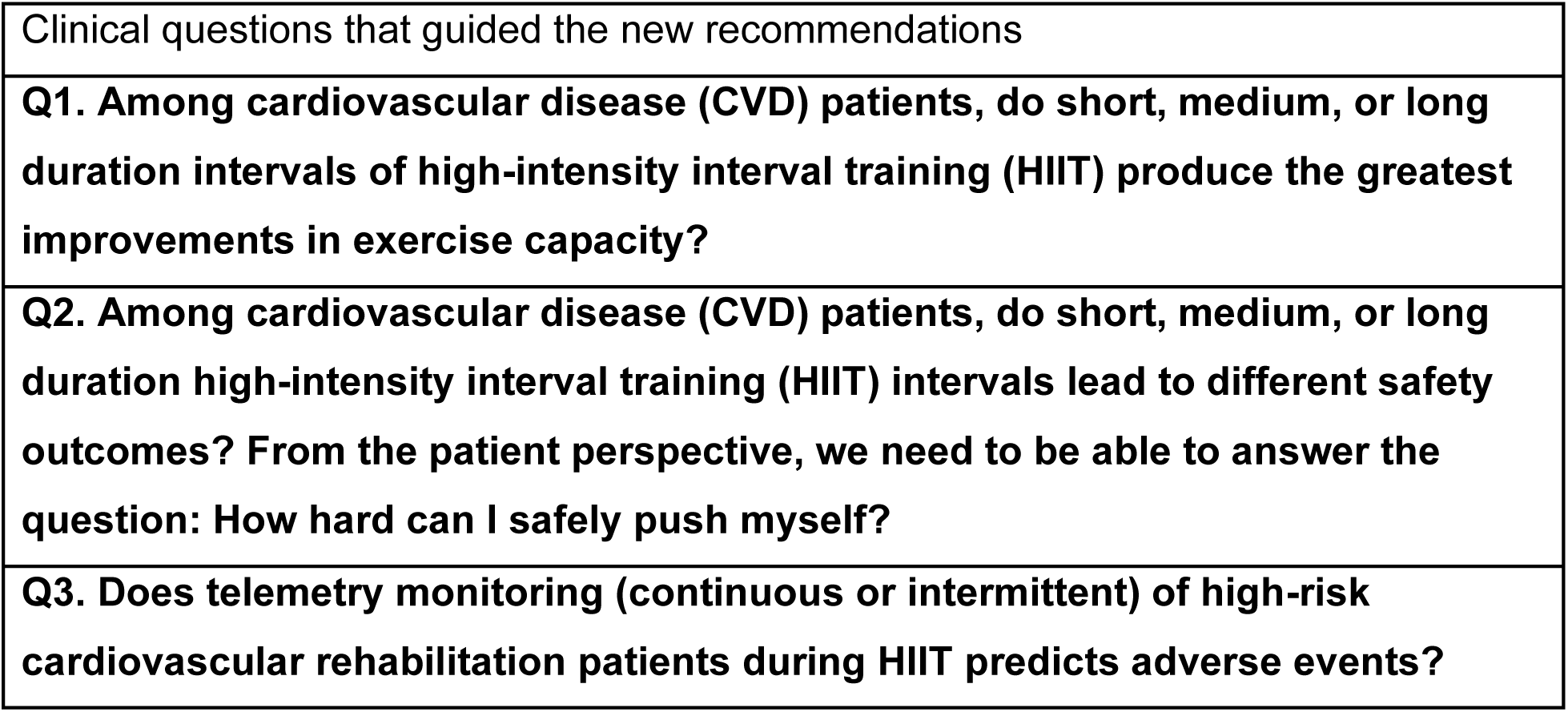

#### 2.5.2 Literature Review - Rapid Review on HIIT

The rapid review was developed according to WHO Practical Guide for Rapid Review26 and reported according to the Preferred Reporting Items for Systematic Reviews and Meta-Analyses (PRISMA).22

#### 2.5.3 Quality assessment

Quality assessment of the studies was conducted using the Tool for the assEssment of Study qualiTy and reporting in EXercise (TESTEX).^27^ In summary, TESTEX is a tool developed to analyze exercise interventions regardless of study design. This tool includes 5 points for study quality and 10 points for reporting and assesses specific quality issues related to the delivery of exercise training interventions. The studies were classified as lower quality (≤7 points) or higher quality (≥ 8 points). Details about the scoring of items are described in the Supplement (Table S3).

#### 2.5.4 Meta-analyses

Exercise capacity data was extracted as described by the authors and transformed when needed (e.g., METS or VO_2max_) to standardise to ml/min/kg for quantitative summary and meta-analysis. To be able to compare different HIIT prescriptions, groups were created based on the active/higher intensity exercise intervals: long duration intervals (HIIT Long; 3 min and higher); and short to medium duration intervals (HIIT Short; 10 sec to 2:59 min). Further, studies that reported having a progressive or mixed prescription loads (e.g., short and long length of intervals in the same week, or across time, for example) were clustered in a “progressive” group, considering they used different intervals along the training period. Mixed-effects models were used for overall effects and to compare groups (i.e., exercise prescriptions and modalities, length of interval training, patients’ profiles). Standardised Mean Difference (SMD), with 95% confidence intervals (CI), between pre- and post-interventions were calculated. According to Cohen’s recommendation^28^, SMDs between 0.2 and 0.5 were considered as a small effect size, between 0.5 and 0.8 moderate, and 0.8 and above as a large effect. Statistical heterogeneity was explored using the I^2^ test and sensitivity analysis (i.e., exploring different HIIT prescriptions and according to quality scoring).

## RESULTS

### 3.1 Systematic review of previous guidance documents

The systematic review identified 59 potential recommendation documents, with only 15 based on a systematic review of the literature, resulting in the inclusion of 15 documents. The list and characteristics of these documents are available in the Supplement (Table S2) and in the Guidelines Report (https://osf.io/9ykrp). The recommendations from these documents were distilled into 9 recommendations. The distillation process was complex due to the different ways in which recommendations were created (i.e., not always based on a PICO), evaluated, graded, and reported. As such, we considered only recommendations based on evidence specifying the populations and outcomes. We then clustered similar recommendations together and extracted evidenced-informed components to create omnibus recommendations. (see https://osf.io/9ykrp for further details)

### 3.2 Recommendations from previous guidance documents

#### 3.2.1 Exercise Assessment

*Synthesis:* Scientific statements and guidelines^29,30^ point to the importance of exercise testing. They draw from cross-sectional and cohort studies that show that parameters evaluated during an exercise test (e.g., blood pressure response, heart rate curves and recovery) were associated with diagnosis,^31–37^ prognosis,^33,38–60^ and exercise capacity. To evaluate these parameters, a graded exercise test may need to be paired with an electrocardiogram.^56,61,62^

***Recommendation 1:*** A supervised graded exercise test conducted as part of the initial cardiovascular rehabilitation assessment will provide information on exercise capacity (e.g., METs on the graded exercise test), diagnosis (e.g., presence of obstructive disease) and prognosis (e.g., risk of cardiovascular events and mortality) (*GRADE: C*).

*Synthesis:* One systematic review showed that the 6-minute walk test can determine functional capacity and correlates with V̇O2 in patients with heart failure.^63^

***Recommendation 2***: For patients with heart failure, a 6-minute walk test may be used to evaluate exercise and functional capacity (*GRADE: B*).

#### 3.2.2 Exercise Prescription – General Patients with CVD

*Synthesis*: Available guidelines^10,11,64^ recommend aerobic exercises with supporting systematic review-level evidence for outcomes such as health-related quality of life, ^13,65–67^ hospital admissions,^65^ and cardiovascular mortality.^2,68–70^. Some documents showed the benefits of resistance exercise as an adjunct to aerobic exercise.^71,72^

***Recommendation 3***: Patients with CVD should be provided with an exercise prescription of aerobic exercise (with or without resistance exercise) to improve general and health-related quality of life, reduce hospital readmissions and, cardiovascular mortality (*GRADE: A*).

*Synthesis*: Available guidelines recommend aerobic exercise to provide benefits for patients with a cardiovascular disease in different settings, such as in hospitals, clinics, and at home. ^73,74,75^ The benefits seemed to be similar in terms of effectiveness and safety.^76–81^

***Recommendation 4***: Aerobic exercise of moderate to vigorous intensity (70-85% peak HR or maximum HR, “moderate” to “somewhat hard” on the Borg Rating Perceived Exertion scale^82^, or 70% V̇O2max) can be prescribed to be completed at home, in the community, or in a hospital setting, with each providing similar effectiveness for improving exercise capacity and being equally safe (in terms of exercise-related cardiovascular symptoms or events) (*GRADE: B*).

*Synthesis*: One trial, referred by one guideline,10 showed the benefits of resistance training for muscle strength and endurance.83

***Recommendation 5***: Providing resistance training as an adjunct to aerobic exercise increases muscle strength and endurance (*GRADE: C*), which are key components to improving functional capacity.

#### 3.2.3 : Exercise Prescription – Patients with Heart Failure

*Synthesis*: Available guidelines^11,30^, supported by systematic review and RCT evidence, recommend exercise to improve quality of life and reduce hospitalisations in patients with heart failure. ^65,84,85^ The benefits of exercise to improve functional capacity were positive but with a lower level of supporting evidence.^86–88^

***Recommendation 6:*** All patients with stable New York Heart Association (NYHA) class I-III heart failure should be enrolled in a CR programme that includes exercise training (aerobic exercise with or without resistance training) to improve exercise capacity (*GRADE: B*), heart failure-related hospitalisations (*GRADE: A*), and health-related quality of life (*GRADE: A*).

*Synthesis*: Exercise prescriptions vary, but a small RCT showed that an exercise programme of walking and cycling at 60-80% of heart rate reserve improved heart rate recovery in patients with heart failure,89 which is a prognostic marker of exercise capacity in this population.56,90

***Recommendation 7***: For patients with heart failure, the aerobic exercise prescription should be for whole-body continuous exercise (e.g., brisk walking, jogging, and cycling) at an initial level rate of “moderate” progressing to “hard” on the Borg Rating Perceived Exertion, 65%-85% maximum HR, or 50%-75% of peak V̇O_2_ (*GRADE: B*).

*Synthesis*: For patients with heart failure and an implantable cardioverter-defibrillator and/or cardiac resynchronisation therapy, multiple RCTs supported the benefits of exercise in improving exercise capacity and quality of life without compromising safety.91–95

***Recommendation 8***: Patients with stable heart failure and an implantable cardioverter-defibrillator and/or cardiac resynchronisation therapy (NYHA functional class II or III), should receive supervised aerobic exercise is recommended to improve exercise capacity and health-related quality of life without increasing the risk of shocks or adverse CVD events (*GRADE: A*).

#### 3.2.4 : Exercise Prescription – Other Populations (65+ years old)

*Synthesis*: Several RCTs have demonstrated the benefits of regular resistance, flexibility, balance, and coordination exercise to improve physical performance in older patients (65+ years old) who have undergone cardiovascular surgery, when combined with aerobic exercise.96,97

***Recommendation 9***: For older patients (65+ years old) who have undergone cardiovascular surgery, resistance, flexibility, balance and/or coordination exercise in combination with aerobic exercise is effective for improving functional capacity (*GRADE: A*).

### 3.3 Newly developed recommendations

#### 3.3.1 Rapid review results

The search was run on July 31, 2023, and yielded 1,338 documents. After duplicates were removed, 1,222 documents were screened for eligibility, and 326 were evaluated for full-text. Of these, 81 original studies were included in the qualitative synthesis (see Supplement, Table S4). We identified seven main clusters of patients with the following conditions:

1. Coronary artery diseases (including coronary artery disease (CAD), myocardial infarction (MI), acute coronary syndrome (ACS), coronary heart disease (CHD), stable angina undergoing percutaneous coronary intervention);
2. Heart failure (HF);
3. Atrial fibrillation (AF);
4. Heart transplant recipients (HTx);
5. Congenital heart disease (ConHD);
6. General CVD (i.e., mixed populations including a variety of patients profiles, such as coronary artery disease, arrhythmias, valvular disease, heart failure, etc.); and
7. Angina with normal coronary arteries (ANOCA)

Table 1 below describes studies’ characteristics according to analysis groups: HIIT groups (all together); HIIT long; HIIT short; HIIT progressive; moderate-to-vigorous intensity continuous training (MICT); control/usual care; and others – according to clinical profile. (see https://osf.io/73g4w/ for more details). There were no HIIT progressive prescriptions in the included heart failure studies. Meta-analyses were conducted using results from CAD, HF, and AF studies only, due to the low number of interventions found for other population groups. Five CAD studies ^98–102^ and three HF studies ^103–105^ were excluded from the meta-analysis, due to the different exercise capacity outcomes (i.e., not using VO_2max_ or METS) or not presenting the data in a way that could be used in the meta-analysis. The remaining seven studies ^106–112^ from other clinical profiles that provided exercise capacity results are described in Table 2.

**Table 1.** Characteristics of studies included in the meta-analyses according to prescription groups.

| Intervention<br>(based on<br>exercise<br>prescription) | Included<br>studies | Total<br>sample<br>(N of<br>patients) | Duration of the<br>intervention<br>(weeks) | Summarised<br>effect on exercise<br>capacity<br>(SMD - mixed-effects analysis) | Exercise<br>intensity (Range) | Reported<br>warm-up<br>(N of studies<br>and range in<br>minutes) | Reported<br>cool-down<br>(N of studies<br>and range in<br>minutes) | Reported<br>familiarisation<br>period<br>(N of studies) |
| --- | --- | --- | --- | --- | --- | --- | --- | --- |
| **Coronary Artery Disease** |  |  |  |  |  |  |  |  |
| **HIIT (short, long**<br>**and progressive)** | Total of studies<br>= 28<br>Number of arms<br>= 32<br>RCT = 22<br>Pre and post =<br>5<br>Retrospective =<br>1 | Total =<br>1,056<br>CAD =<br>527<br>CHD =<br>163<br>MI = 128<br>MI+PCI =<br>218<br>PCI = 20 | Mean: 13.36<br>Median: 12<br>Range: 8-52 | 0.696 (0.592-0.801) | VO2peak: 70–<br>100%<br>HRpeak: 70–<br>100%<br>HRR: 80-95%<br>PPO: 75–121%<br>RPE: 12–18<br>100% of the<br>average maximum<br>workload<br>50% of the<br>maximum load<br>reached with the<br>SRT<br>(peak intervals) | 23<br>(2-15 min) | 19<br>(3-13 min) | 13 |
| **HIIT short** | Total of studies<br>= 8<br>RCT = 6<br>Pre and post =<br>2 | Total =<br>325<br>CAD =<br>100<br>CHD = 12<br>MI = 18 | Mean: 10.5<br>Median: 8<br>Range: 8-16 | 0.637 (0.449-0.825) | PPO: 80-121%<br>VO2peak: 70–<br>100%<br>RPE: 14–17<br>50% of the<br>maximum load | 8<br>(5-15 min) | 8<br>(5-13 min) | 8 |
|  |  | MI+PCI = 195 |  |  | reached with the SRT<br>(peak intervals)<br>HRpeak: 90% |  |  |  |
| **HIIT long** | Total of studies = 16<br>Number of arms = 20<br>RCT = 13<br>Pre and post = 3 | Total = 574<br>CAD = 312<br>CHD = 151<br>MI = 68<br>MI+PCI = 23<br>PCI = 20 | Mean: 14.78<br>Median: 12<br>Range: 8-52 | 0.589 (0.520-0.658) | VO2max: 60-90%<br>HRpeak: 80-95%<br>HRmax: 80-100%<br>HRR: 80-90%<br>RPE: 15-18 | 14<br>(2-10 min) | 11<br>(3-5 min) | 4 |
| **HIIT progressive** | Total of studies = 4<br>RCT = 3<br>Retrospective = 1 | Total = 157<br>CAD = 115<br>MI = 42 | Mean: 12<br>Median: 12<br>Range: 12 | 1.274 (0.784-1.764) | HRpeak: 90%<br>HRR: 85-95%<br>RPE: 15-17<br>Average maximum workload: 100% | 3<br>(5-10 min) | 2<br>(4-10 min) | 1 |
| **MICT** | Total of studies = 16<br>RCT = 14<br>Pre and post = 1<br>Retrospective = 1 | Total = 543<br>CAD = 302<br>MI = 33<br>MI+PCI = 208 | Mean: 14.60<br>Median: 12<br>Range: 8-52 | 0.463 (0.314-0.612) | VO2peak: 50-80%<br>HRpeak: 60-75%<br>HRR: 40-80%<br>PPO: 51-65%<br>RPE: 11-14<br>HR at VT1 +10% | 12<br>(5-15 min) | 11<br>(5-13 min) | 6 |
|  |  |  |  |  | Maximal exercise capacity: 60-80% |  |  |  |
| **Control or usual care** | Total of studies = 6 RCT | Total = 171<br>CAD = 130<br>CHD = 11<br>MI = 10<br>PCI = 20 | Mean: 16.33<br>Median: 14<br>Range: 8-26 | -0.007 (-0.259-0.245) | NA | NA | NA | NA |
| **Heart Failure** |  |  |  |  |  |  |  |  |
| **HIIT (short and long)** | Total of studies = 28<br>Number of arms = 31<br>RCT = 23<br>Pre and post = 5 | Total = 742<br>HF = 91<br>HFpEF = 122<br>HFrEF = 529 | Mean: 14.87<br>Median: 12<br>Range: 12-52 | 0.721 (0.559-0.884) | VO2peak: 80-95%<br>VO2reserve: 80-90%<br>HRpeak: 85-95%<br>HRR: 75-90%<br>HRmax: 90-95%<br>RPE:: 15-17<br>WRpeak: 80-100%<br>SRT: 50% workload<br>HR range: 5% to 10% above VT1 | 23<br>(1-15 min) | 21<br>(1-15 min) | 9 |
| **HIIT short** | Total of studies = 5 RCT | Total = 76<br>HF = 21<br>HFrEF = 55 | Mean: 14.40<br>Median: 12<br>Range: 12-24 | 0.484 (0.209 – 0.758) | WRpeak: 80-100%<br>SRT: 50% workload | 1 (NR) | 1 (NR) | 3 |
| **HIIT long** | Total of studies = 23<br>Number of arms = 26<br>RCT = 18<br>Pre and post = 5 | Total = 666<br>HF = 70<br>HFpEF = 122<br>HFrEF = 474 | Mean: 14.96<br>Median: 12<br>Range: 12-52 | 0.779 (0.594 – 0.964) | VO2peak: 80%-95%<br>VO2reserve: 80-90%<br>HRpeak: 85-95%<br>HRR: 75-90%<br>HRmax: 90-95%<br>RPE: 15-17<br>WRpeak: 90%<br>HR range: 5% to 10% above VT1 | 22<br>(1-15 min) | 20<br>(1-15 min) | 6 |
| **MICT** | Total of studies = 11 RCT | Total = 289<br>HF = 37<br>HFpEF = 63<br>HFrEF = 189 | Mean: 16.73<br>Median: 12<br>Range: 12-52 | 0.297 (0.136-0.458) | WRpeak: 50% workload<br>HRpeak: 60-75%<br>HRR: 35-60 %<br>VO2peak: 40-60%<br>VO2reserve: 50-60% | 4<br>(3-10 min) | 4<br>(3-10 min) | 4 |
| **Control or usual care** | Total of studies = 16<br>Number of arms = 18<br>RCT = 14 | Total = 450<br>HF = 49<br>HFpEF = 52 | Mean: 15.18<br>Median: 12<br>Range: 12-52 | -0.100 (-0.220-0.021) | NA | NA | NA | NA |
|  | Pre and post = 2 | HFrEF = 349 |  |  |  |  |  |  |
| **Combined** | Total of study = 3 RCT | Total = 46<br>HF = 25<br>HFrEF = 21 | Mean: 12<br>Median: 12<br>Range: 12 | 0.382 (0.150-0.614) | VO<sub>2</sub>peak: 80%<br>50% workload<br>steep ramp test (SRT) | 2**<br>(3 min) | 2**<br>(3 min) | 1 |
| **Atrial Fibrillation** |  |  |  |  |  |  |  |  |
| **HIIT** | Total of studies = 4* RCT | Total = 91<br>AF = 84<br>AF + HFrEF = 7 | Mean: 25.5<br>Median: 19<br>Range: 12-52 | 1.059 (0.477-1.642) | HRpeak: 85%-95%<br>Borg: 16–18 (80% of perceived exertion) | 4<br>(5-10 min) | 4<br>(5-10 min) | 2 |
| **Control or usual care** | Total of studies = 3* RCT | Total = 64<br>AF = 55<br>AF + HFrEF = 9 | Mean: 30<br>Median: 26<br>Range: 12-52 | 0.287 (0.496-1.070) | NA | NA | NA | NA |

\*One of the study presents data for a population with both AF and HFrEF
\*\*One did not report the duration of the warm up or cool down
†Borg scale: 6-20

**Table 2.**
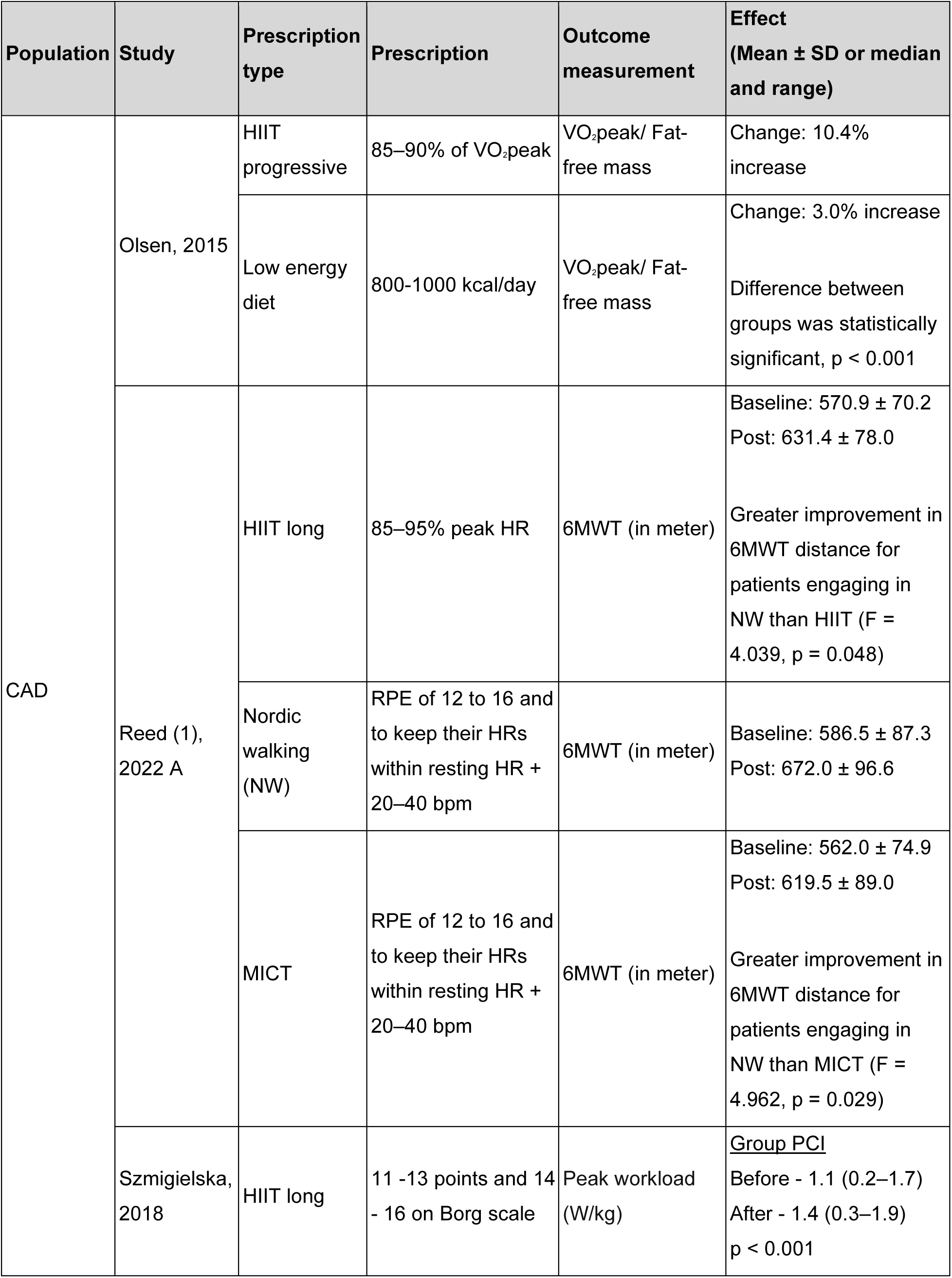
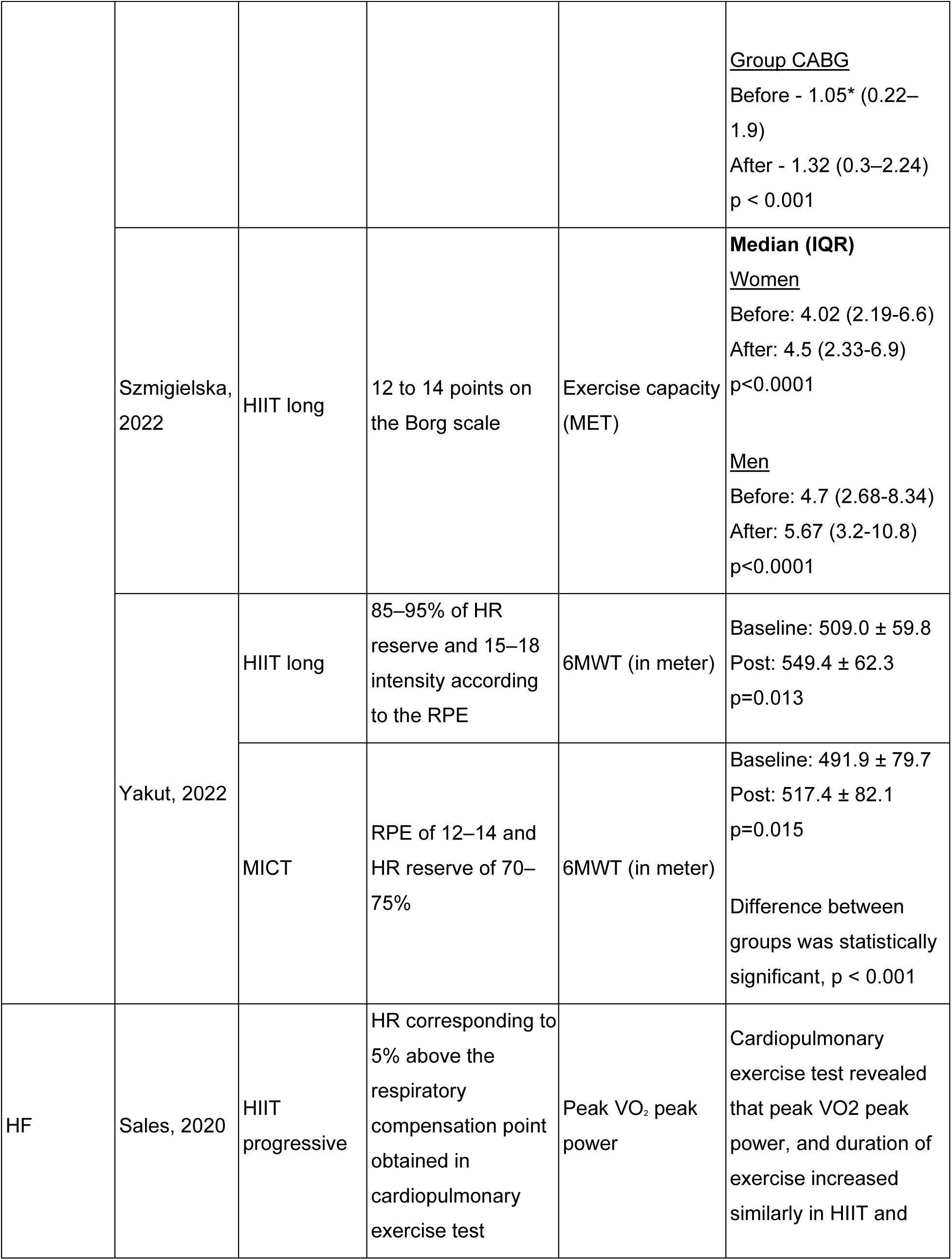
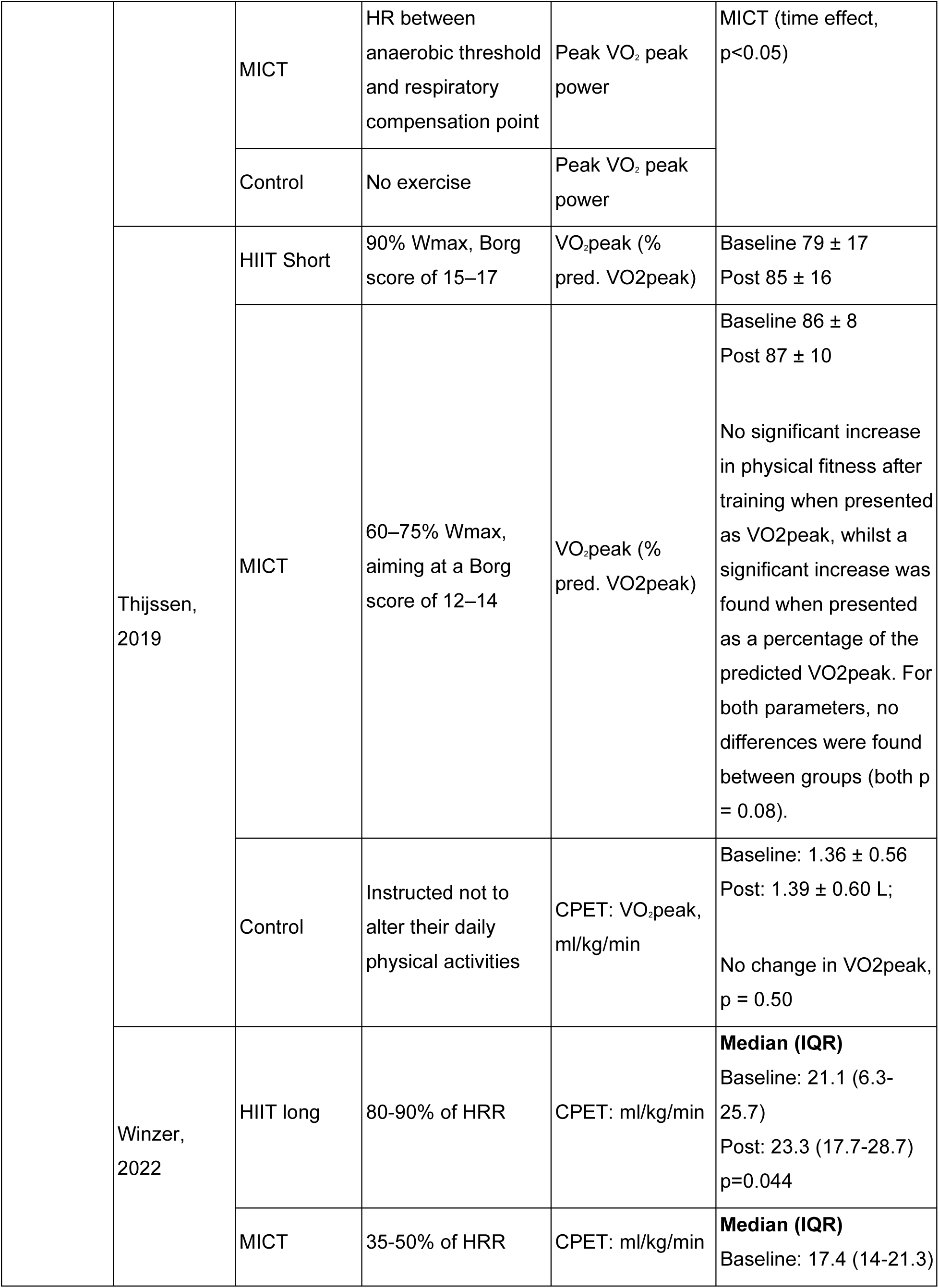
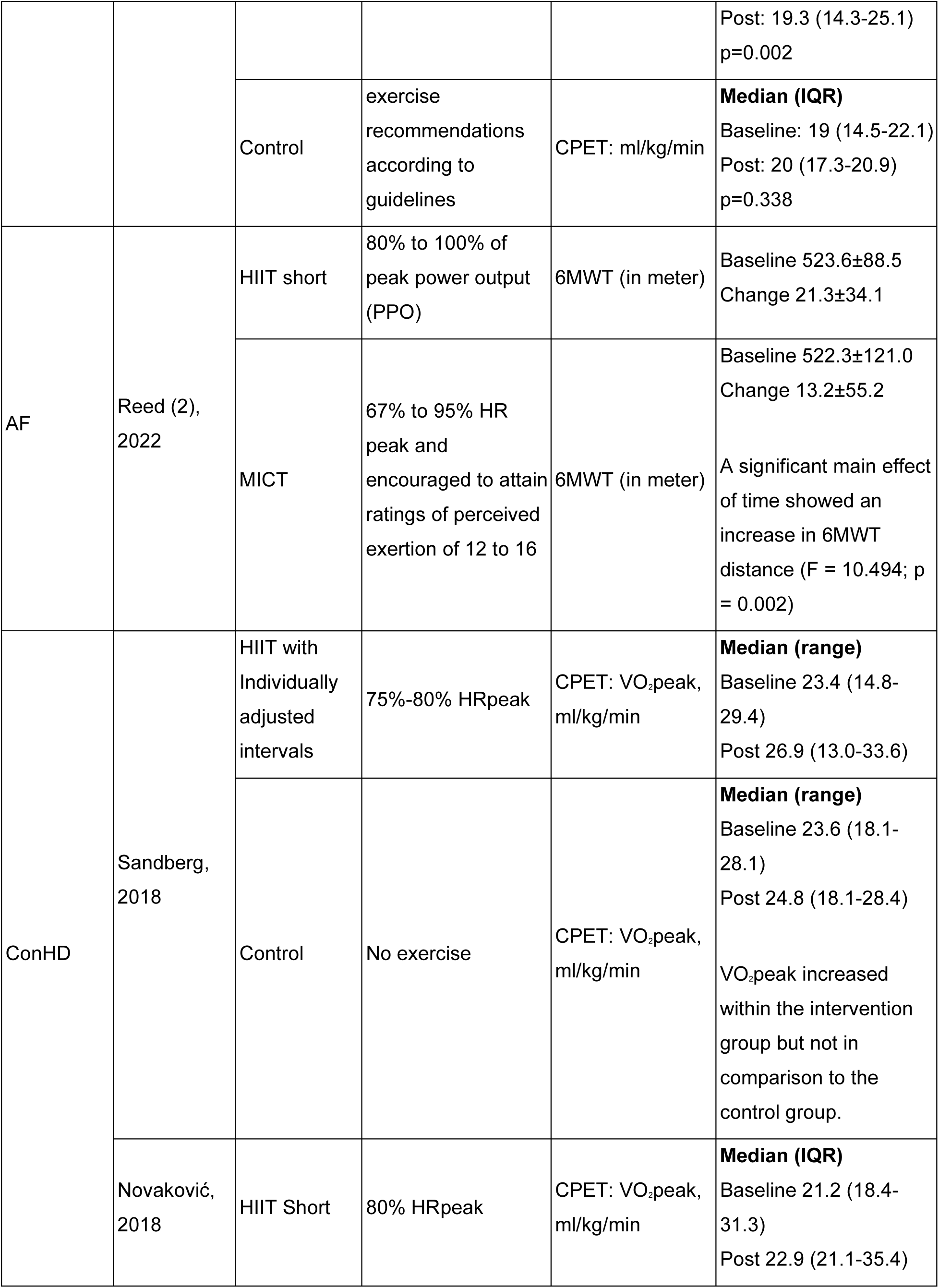
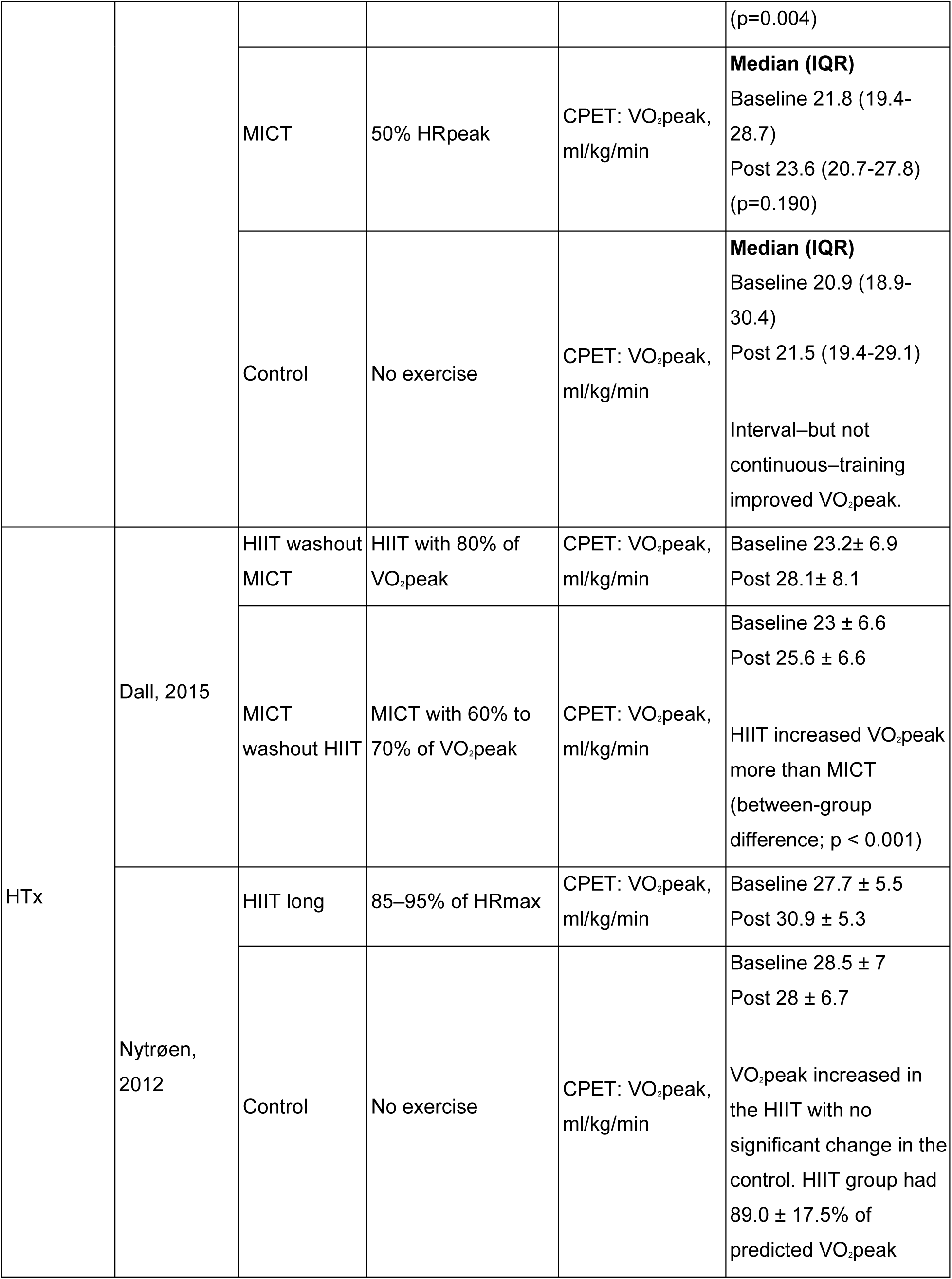
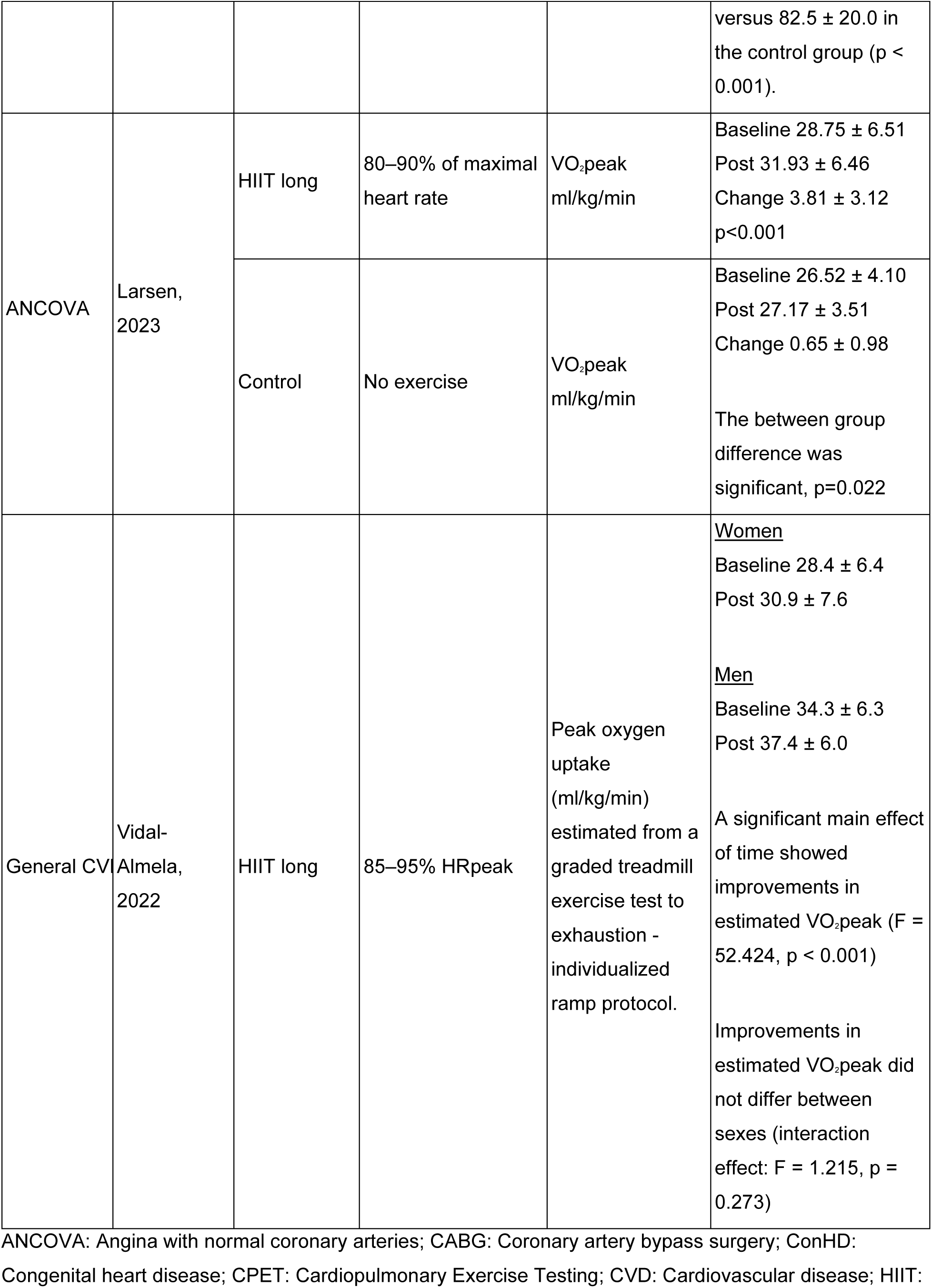
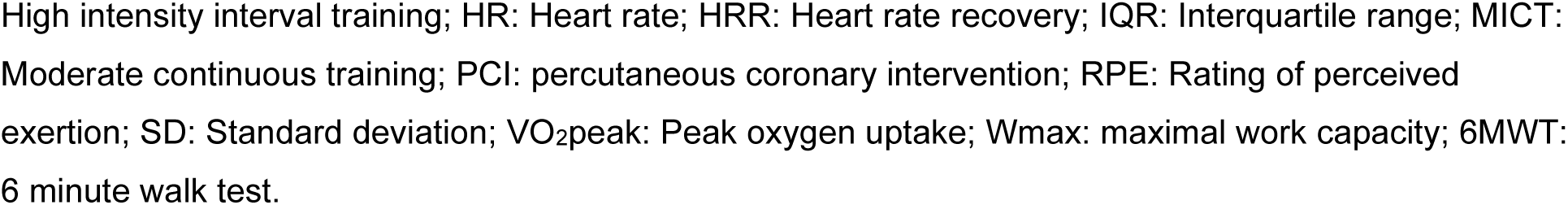
Exercise capacity results of the studies not included in the meta-analysis.

#### 3.3.2 HIIT effect on Exercise Capacity (VO2max) for patients with CAD: HIIT vs MICT

*Synthesis:* From table 3, there is evidence that HIIT is both statistically and clinically superior to MICT for exercise capacity improvement in patients with CAD, when directly compared within studies.

**Table 3.** Summary from the meta-analysis results for direct comparisons of HIIT vs. MICT on changes in exercise capacity in patients with CAD.

| Population | Intervention | Point estimate (IC) Difference in mean exercise capacity change (VO<sub>2max</sub>- ml/min/kg) | N of arms |
| --- | --- | --- | --- |
| CAD | Direct Comparison p= 0.003; I<sup>2</sup> = 89.11% |  |  |
|  | HIIT | 4.54 (3.44 – 5.64) | 16 |
|  | MICT | 2.50 (1.71 – 3.30) | 16 |

CAD: coronary artery disease; HIIT: High intensity interval training; MICT: Moderate intensity continuous training; VO<sub>2max</sub>: Maximum rate of oxygen consumption

**Recommendation 10**: Patients with known CAD should be considered for a HIIT prescription to optimise improvements in exercise capacity (Grade B).

#### 3.3.3 HIIT effect on Exercise Capacity (VO2max) for patients with HF: HIIT vs MICT

*Synthesis:* From table 4, there is no statistical nor clinical difference in the effects of HIIT or MICT for improving exercise capacity in patients with HF, when directly compared within studies.

**Table 4.** Summary from the meta-analysis results for direct comparisons of HIIT vs. MICT on exercise capacity in patients with HF.

| Population | Intervention | Point estimate (IC) Difference in mean exercise capacity change (VO<sub>2max</sub>- ml/min/kg) | N of arms |
| --- | --- | --- | --- |
| HF | Direct Comparison p=0.122; I<sup>2</sup> = 76.64% |  |  |
|  | HIIT | 2.54 (1.44 – 3.63) | 11 |
|  | MICT | 1.62 (1.25 – 2.00) | 11 |

HF: Heart failure; HIIT: High intensity interval training; MICT: Moderate intensity continuous training; VO<sub>2max</sub>: Maximum rate of oxygen consumption

***Recommendation 11***: Patients with known HF should be considered for either a HIIT or MICT prescription to improve exercise capacity (Grade B).

#### 3.3.4 Effect of HIIT interval duration on Exercise Capacity for patients with CAD or HF

*Synthesis: From table 5,* all durations of HIIT intervals provided a positive benefit for exercise capacity in patients with known CAD or HF. For CAD, progressive HIIT (HIIT protocol using different interval lengths along the training protocol) seemed to provide greater benefits but more research is needed as only four studies used this prescription. ^113–116^ For HF, no difference was detected between different interval lengths.

**Table 5.**
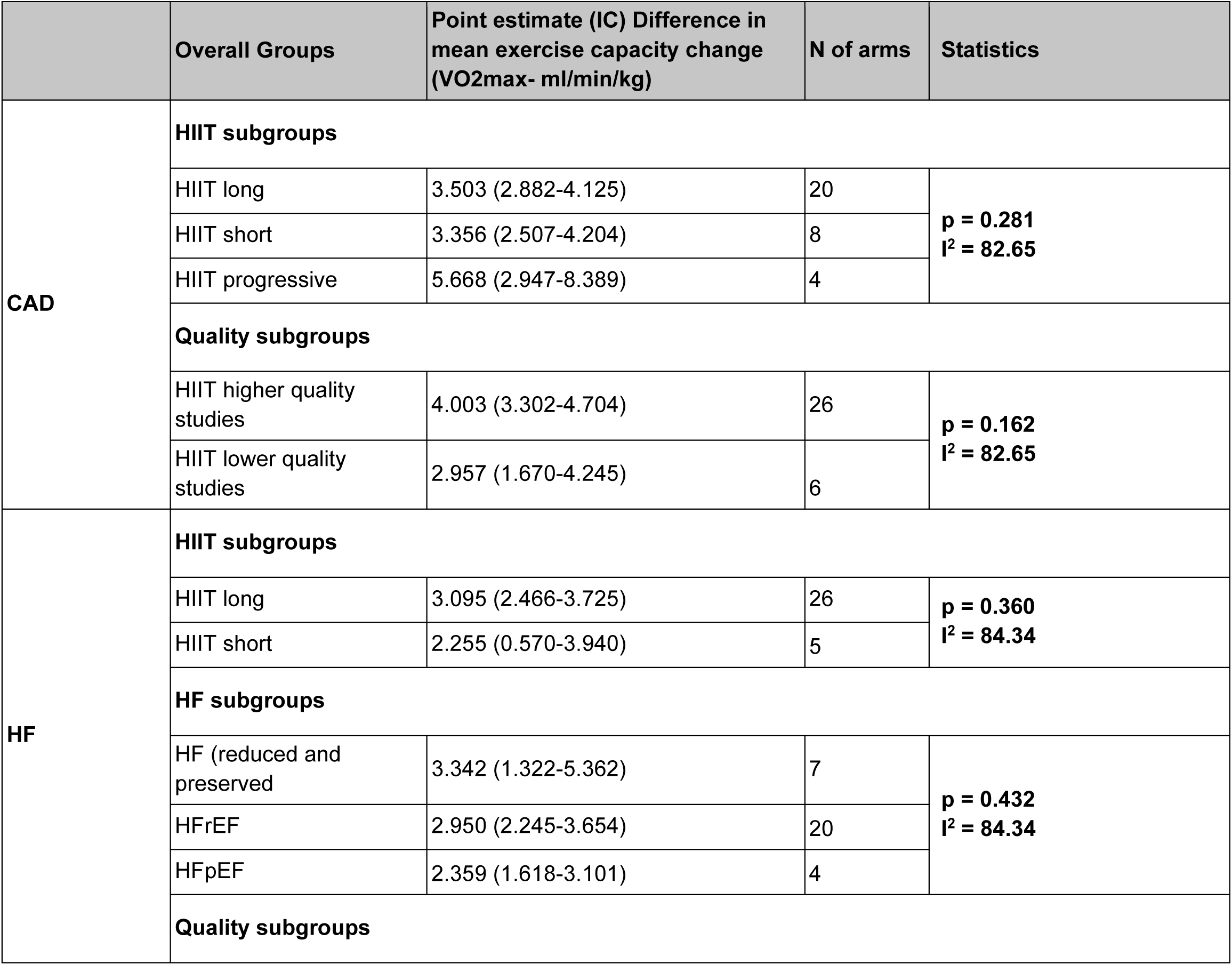
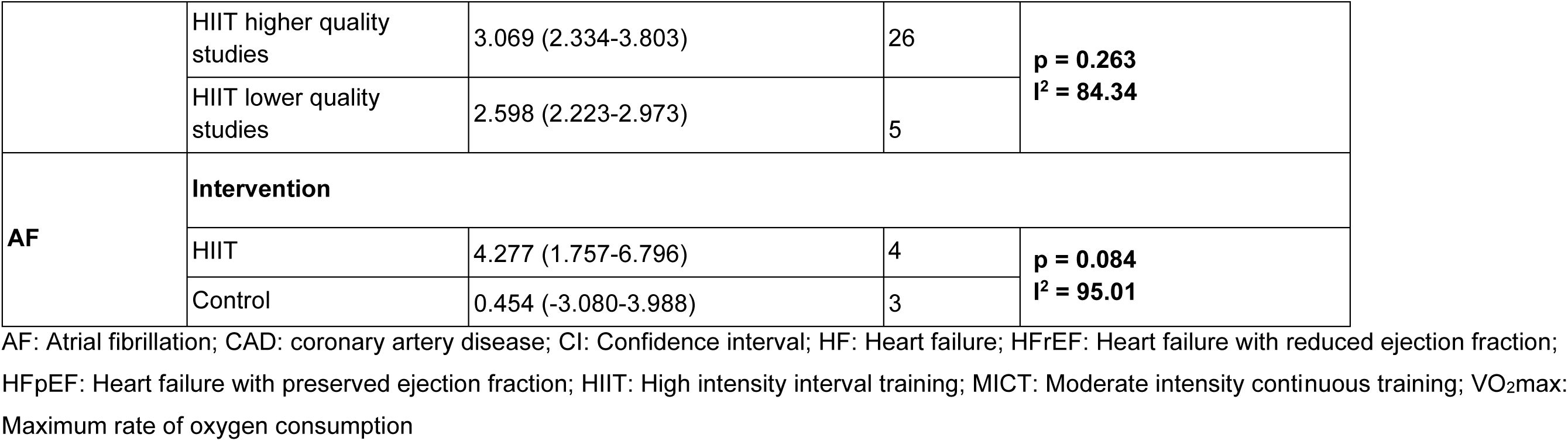
Results of the meta-analysis on the impact of HIIT on exercise capacity.

***Recommendation 11:*** In patients with known CAD or HF who partake in HIIT, exercise prescriptions may employ any duration of high-intensity intervals to improve exercise capacity (*Grade B*).

### 3.4 Insights from the rapid review for future research

#### 3.4.1 Effect of HIIT on Exercise Capacity (VO2max) for patients with atrial fibrillation: exercise vs control

*Synthesis:* From the individual interventional arms, HIIT showed improvements in exercise capacity for individuals with atrial fibrillation, with no improvement seen within the control arms. However, due to the fact that only 4 studies were included ^117–120^, there was only a non-statistically significant trend for a difference between the arms (p=0.084). This means that while there may be indications suggesting the potential superiority of HIIT over control, the available data does not provide conclusive evidence in its favour.

***Recommendation for Future Research 1 (PICO 1):*** More and larger studies are needed exploring the impact HIIT, relative to an appropriate comparison group, on exercise capacity in individuals with atrial fibrillation.

#### 3.4.2 The safety of HIIT

*Synthesis*: There was no evidence of any notable adverse events in response to HIIT (see Supplement, Table S5 and https://osf.io/4uwa5). However, this conclusion is tempered by the fact that most studies did not systematically record or report adverse events, and for those that did, there was no consistent reporting standard used. This means that more data and consistent reporting are needed to be able to comment on the safety of HIIT. The use of tools such as the CACPR Registry could be considered for future studies.

***Recommendation for Future Research 2 (PICO 2):*** The safety of HIIT in CR is still unclear and needs further exploration. There is a critical need to have consistent standardised approaches in reporting of both efficacy and safety outcomes for HIIT in CR.

#### 3.4.3 The use of telemonitoring in the context of HIIT

*Synthesis:* There was sparse research exploring the use of telemetry and prescribing HIIT in patients with CAD or HF. No studies investigated or addressed telemonitoring or issues of safety in these populations (Supplement Table S5). Given the greater emphasis on telemedicine since the start of the COVID-19 pandemic, understanding the benefits and potential negative outcomes is critical in being able to optimise both remote interventions and monitoring.

***Recommendation for Future Research 3 (PICO 3):*** The use of telemonitoring in the context of HIIT in CR needs to be explored.

#### 3.4.4 The role of HIIT in CR populations other than those with CAD or HF

*Synthesis*: Most research was conducted in patients with CAD or HF. With the increase in more diverse pathophysiology coming to CR (e.g. AF, heart transplant valvular disease, spontaneous coronary artery dissection, peripheral artery disease), there is a need to study the efficacy of HIIT used for the management of other cardiovascular populations.

***Recommendation for Future Research 4:*** The role of HIIT in CR populations other than those with CAD or HF (e.g., AF, heart transplant, etc.) needs to be explored further.

#### 3.4.5 The optimal HIIT protocol

*Synthesis*: Though we were able to explore the question of HIIT interval length there were several parameters (e.g., frequency, type, duration, inclusion of warm-up and cool-down) key to the HIIT prescription that we were not able to explore due to a lack of data. This will be critical to providing detailed prescriptions to specific populations. In addition, greater consistency in the use of intensity determinants (e.g., %PPO, % V̇O_2peak_, %HR_max_, RPE) for investigating HIIT prescription options and their respective outcomes is needed.

***Recommendation for Future Research 5:*** More studies directly exploring and undertaking comparative analyses of the various HIIT protocols, (e.g., intensity of the interval, intensity of the recovery interval, the number of intervals per session, etc.), are needed to identify the optimal prescription for various CR populations.

## 4. Considerations in the interpretation of the available evidence

While exercise recommendations within the context of CR have seen significant advancements, there remains a great deal of inconsistency across different guidelines. Most notably, very few follow a systematic process to synthesize the extant literature to form recommendation which made it challenging to identify consistencies across documents. However, when a consensus did exist, there was substantial supporting literature. For the purpose of the current document, we only considered recommendations based on evidence specifying the disease population and outcomes.

While the overall quality of the studies included in the rapid review was good, with an average score of 9.5 out of 15 the heterogeneity of the reporting made assessment challenging (20 studies scoring above 12, and 15 scoring 7 or less). This issue was particularly evident in the reporting of adverse events (AE), as the few studies that did report AE varied in their time of assessment (e.g, during the exercise training, during the testing, during the study period) and classification methods (i.e., definition of seriousness, reporting if the event was related or non-related to exercise). This lack of consistent information meant that AEs were only reported qualitatively, specifying the time of assessment when possible. Additionally, the number of available studies with HIIT prescriptions was quite heterogeneous leading to a lack of studies that had direct comparisons. This challenges the ability to properly compare variations in interval length. We aimed to categorise the HIIT prescriptions into four groups (i.e., HIIT short, medium, long, progressive or mixed lengths – overall 5 groups) for the meta-analysis sensitivity analyses, but had to reclassify them to short, long, and progressive only because of the number of studies available for some of the groups.

## 5. Conclusion

Overall, the present CACPR guidelines and guidelines from international associations consistently recommend aerobic exercise for CR. This document highlights its potential benefits for patients with CVD, including improved exercise capacity and reduced hospitalisation and mortality. The review showed that various HIIT interval lengths improved exercise capacity in individuals with CAD or HF. These recommendations were created based on the evidence syntheses generated to support the working group’s decisions. Since then, with the continuous growth of literature in this area, additional studies have been published. Nonetheless, further research is required to refine and expand these recommendations and explore the safety aspects of these interventions with more rigorous methodologies. Further research is also needed to assess if specific HIIT interval lengths could have a greater effect on exercise capacity and to include other populations, such as those with AF or broader CVD profiles. Transparent and ongoing research efforts are essential for developing clearer and more comprehensive guidelines in the future.

## Data Availability

All data produced in the present study are available upon reasonable request to the authors.

https://osf.io/w2kcv/

## Acknowledgments

This document’s successful completion is the result of collaborative efforts. We would like to thank everyone involved. We would also like to acknowledge the participation of additional researchers who contributed to pieces of the evidence synthesis reports, including study selection, data extraction and report development: Dr David Anekwe, Dr Jovana Stojanovic, Laurence Paquet, and Doro Yip.

## Contributions

Decisions around the project management and selection of the working group members: CACPR Executive Committee for Guidelines (Ariany Marques Vieira, Ashlay Huitema, Diana Hopkins-Rosseel, Jennifer Reed, Lisa Cotie, Neville Suskin, Nicola Paine, Paula Aver Bretanha Ribeiro, Paul Oh, Simon L. Bacon)

Project management: Ariany M Vieira, Paula A B Ribeiro, and Simon L Bacon. Evidence synthesis team: Ariany M Vieira, Paula A B Ribeiro, Laurence Paquet, Doro Yep.

Guidance and training of the Working Group: Ariany M Vieira, Paula A B Ribeiro. Moderating the discussions, helping the group move forward with the meeting agendas, managing feedback, and assisting members in related activities: Exercise Working Group Chairs (Diana Hopkins-Rosseel, Jennifer Harris).

Knowledge synthesis interpretation, generating and prioritizing the clinical questions, developing the recommendations: Exercise Working Group Members (Andree-Anne Hebert, Billie-Jo Mabey, Colin Yeung, Diana Hopkins-Rosseel [Chair], Dustin Kimber, Jennifer Harris [co-Chair], Kimberley Way, Nancy Hansen, Patrick Warner, Stephanie Prince Ware, Tim Hartley).

Drafting of the document: Ariany M Vieira, Paula A B Ribeiro, Laurence Paquet and Simon L Bacon.

Project Coordinator and Supervisor: Simon L Bacon.

## Abbreviations and Acronyms

1RM: One-repetition maximum
AE: Adverse event
AF: Atrial fibrillation
CABG: Coronary artery bypass grafting
CACPR: Canadian Association of Cardiovascular Prevention and Rehabilitation
CAD: Coronary artery disease
CCS: Canadian Cardiovascular Society
CHD: Congestive heart disease
ConHD: Congenital heart disease
CPET: Cardiopulmonary Exercise Testing
CR: Cardiovascular rehabilitation
CRT: Cardiac synchronization therapy
CRT-D: Cardiac resynchronization therapy and defibrillation device
CVD: Cardiovascular disease
EWG: Exercise working group
HF: Heart failure
HFpEF: Heart failure with preserved ejection fraction
HFrEF: Heart failure with reduced ejection fraction
HR: Heart rate
HRmax: Maximum heart rate
HRpeak: Peak heart rate
HRR: Heart rate reserve
HIIT: High-intensity interval training
HTx: Heart Transplant
ICD: Implantable cardioverter defibrillator
IMT: inspiratory muscle training
LVAD: Left ventricular assist device
IMW: Inspiratory muscle weakness
MI: Myocardial infarction
MICT: Moderate-intensity continuous training
NA: Not applicable
NR: Not reported
PCI: percutaneous coronary intervention
PICO: Template to describe the clinical question (Population, Intervention, Comparison groups and Outcome)
PPO: Peak power output
PRM: Physical Rehabilitation Medicine
RCT: randomized controlled trial
RPE: Rating of perceived exertion
SMD: Standardized Mean Difference
SRT: Steep ramp test
VO2max: Maximum rate of oxygen consumption
VO2peak: peak oxygen uptake
VO2 reserve: difference between maximum and resting
VO2 VT: Ventilatory threshold
WHO: World Health Organization
WK: Workload
Wmax: maximal work capacity
WRpeak: Peak work rate

