## Supplemental Material for "Canadian Association of Cardiovascular Prevention and Rehabilitation (CACPR) Exercise Training Recommendations in Cardiovascular Rehabilitation"

### CACPR Exercise Guidelines

**Supplement**

#### Overview of the project

CACPR new guidelines development - creating working groups and workflow:

1. Selection of CR Components, the mains topics to be explored by working groups: A survey-based on a Delphi method consulted experts and patients in the field to develop the CR thematic priority list (e.g., nutrition, physical activity, optimisation of cardiovascular drug therapy, general behaviour change program), helping the Executive Committee to establish priorities and define the starting point of the process. A pre-priority list was first created by experts and researchers in CR and was validated by the target population (healthcare professional, patients and other interested and affected parties). Exercise was elected as the highest priority.
2. Recruitment of the Exercise Working groups members: A public call for members was disseminated through CACPR and social media. Recruitment also announced in the Fall Conference 2019, when the project was in the planning stages. Personal invitations to experts were sent by the Executive Committee members. Applications were submitted online via RedCap. The selection of members for the Exercise working group was made by the Executive Committee, and considered a pan-Canadian representation, including Francophones, balanced representation of CR roles and career stages, and equal representation of men and women. The group included 11 members, with two patients and carers representatives. It also included researchers and healthcare professionals with a variety of roles within CR (general practitioners, public health physicians, nurses, psychologists and others).
3. The systematic review of literature - previous CR guidelines: this phase was used to identify the high-quality extant exercise-based recommendations so as not to replicate prior work.
4. Creation of PICO/PECO statements for systematic searches: the Exercise working group participated in an internal priority setting process with anonymous voting to select the main questions that weren't addressed by current recommendation documents.
5. Rapid review of PICOs: Generation of an evidence summary by META Group to support the exercise working group discussions and evidence-informed decisions.
6. Recommendation development: the evidence reports were received and analyses by the Exercise Working group members who generated the new recommendations which were reviewed by META groups and graded using Hypertension Canada scoring system. Only recommendations with a Grade of A-C were considered.
7. Recommendation validation: the final evidence reports and recommendations were reviewed by the central review committee (CRC) to provide external validation of their appropriateness.

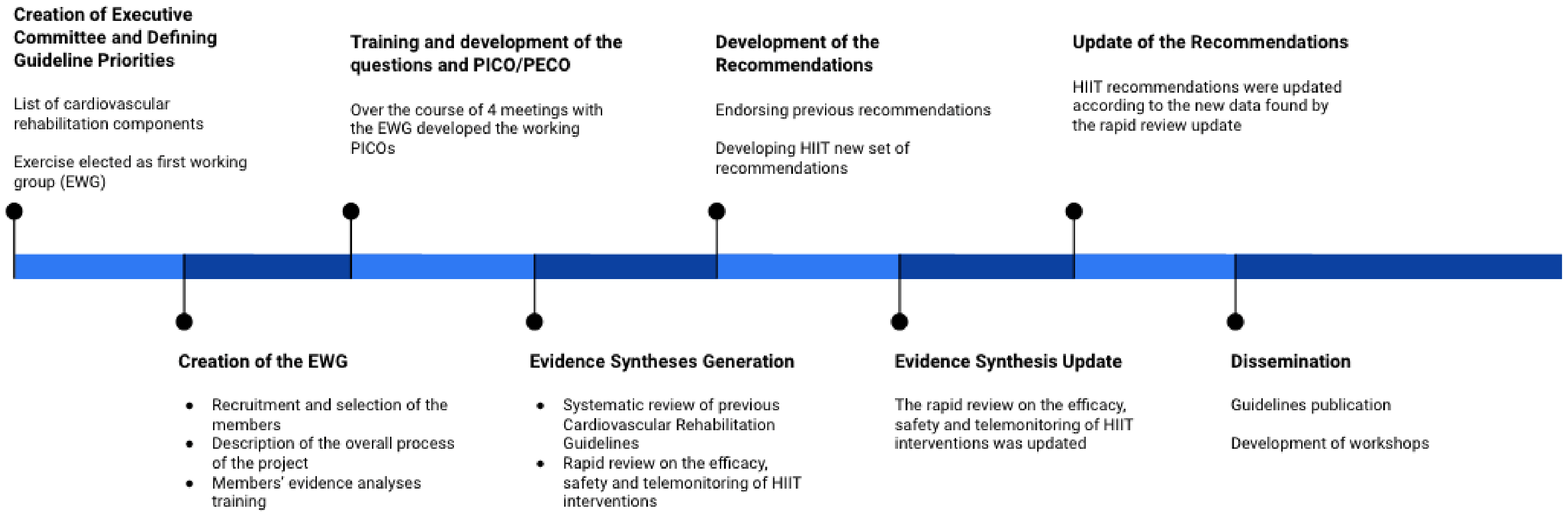

*Figure 1. Timeline of the guideline development*

*Table S1. Exercise working group member description*

| Name | Member description | Province |
|---|---|---|
| Diana Hopkins-Rosseel | Physiotherapist / rehab coordinator / professor | Ontario |
| Jennifer Harris | Physiotherapist - CR coordinator | Ontario |
| Nancy Hansen | Patient | Ontario |
| Tim Hartley | CSEP-CEP/Certified Exercise Physiologist/ Kinesiologis | Ontario |
| Andrée-Anne Hebert | Exercise specialist or Kinesiologist | Québec |
| Dustin Kimber | Kinesiologist - specialist in chronic diseases | Manitoba |
| Billie-Jo Mabey | Nurse | New-Brunswick |
| Stephanie Prince Ware | Researcher | Ontario |
| Patrick Warner | Patient | Ontario |
| Kimberley Way | Exercise physiologist and researcher | Ontario |
| Colin Yeung | Cardiologist | Saskatchewan |

#### Systematic review of exercise cardiac rehabilitation recommendations

*Table S2. List of included documents (N=15)*

| **Author (Year)** | **Title** | **Journal /Endorsement/ Country** | **Type** | **Target Patient Population** | **Stakeholder** | **Other Components (Y/N)** | **Report on COI: (D/N/NR) Funding: (Name/N/NR)** |
|---|---|---|---|---|---|---|---|
| NICE (2020) | Acute coronary syndromes (NG185) | https://www.nice.org.uk/guidance/ng185 National Institute for Health and Care Excellence / England, UK | Guideline | Patients post-acute coronary syndromes (ST-segment elevation myocardial infarction, non-ST-segment elevation myocardial infarction and unstable angina). | HCP | Y | COI: NR<br><br>FUND: NICE |
| Ambrosetti (2020) | Secondary prevention through comprehensive cardiovascular rehabilitation: From knowledge to implementation. 2020 update. A position paper from the Secondary Prevention and Rehabilitation Section of the European Association of Preventive Cardiology | European Journal of Preventive Cardiology / European Association of Preventive Cardiology / European countries | Position paper | Various clinical conditions: post-acute coronary syndrome and post-primary coronary angioplasty, chronic coronary syndromes, cardiac surgery – coronary artery or valve heart surgery, chronic heart failure, cardiac transplantation, ventricular assist devices, peripheral artery disease. | CR professionals | Y | COI: N<br><br>FUND: N |
| Thomas (2019) | Home-based cardiac rehabilitation: a scientific statement from the American Association of Cardiovascular and Pulmonary Rehabilitation, the American Heart | Circulation / American Heart Association, and the American College of Cardiology / USA | Scientific statement | CR patients | NR *(Policymakers and third-party payers)* | Y | COI: D<br><br>FUND: NR |

| | | | | | | | |
|---|---|---|---|---|---|---|---|
| | Association, and the American College of Cardiology | | | | | | |
| Lugo-Agudelo (2019) | Clinical Practice Guidelines for the prevention, diagnosis, treatment, and rehabilitation of heart failure in a population greater than 18 years-old: B, C and D Classification. Cardiac rehabilitation component | Revista Colombiana de Cardiologia / Sociedad Colombiana de Cardiologıa y Cirugıa Cardiovascular / Columbia | Clinical Practice Guideline | Heart failure (mainly) and patients with cardiac resynchronization therapy, implantable cardio-defibrillator and implantable cardio-defibrillator cardiac resynchronization therapy | CR professionals | N | COI: N<br><br>FUND: El desarrollo de esta guía fue financiado por el Ministeriode Salud y Protección Social, por medio de la convocato-ria |
| Juocevicius (2018) | Evidence-based position paper on Physical and Rehabilitation Medicine (PRM) professional practice for people with cardiovascular conditions. The European PRM position (UEMS PRM Section) | European journal of physical and rehabilitation medicine / European Union through the European Union of Medical Specialists (UEMS) of Physical and Rehabilitation Medicine / European countries | Position paper | People with cardiovascular conditions | Physicians | Y | COI: N<br><br>FUND: NR |
| Scottish Intercollegiate Guidelines Network (2017) | Guideline 150: Cardiac rehabilitation - Full guideline | www.sign.ac.uk / Healthcare / Scotland, UK | Guideline | Patients with cardiac disease | Cardiologists, physicians, dietitians, exercise professionals, general practitioners, health service managers, primary and secondary care nurses, | Y | COI: D<br><br>FUND: NR |

| | | | | | | | |
|---|---|---|---|---|---|---|---|
| | | | | | occupational therapists, pharmacists, physiotherapists, psychologists, specialist nurses, academics, third-sector organisations and other healthcare professionals working with patients with cardiac disease | | |
| Grace (2016) | Cardiac rehabilitation delivery model for low-resource settings | Heart / International Council of Cardiovascular Prevention and Rehabilitation and many other national associations / Low-income countries | Consensus statement | All subjects recovering from major coronary heart disease events | CR can be adapted by delivery by non-physician healthcare workers, in non-clinical settings | Y | COI: N<br><br>FUND: N |
| Achttien 2015) | Exercise-based cardiac rehabilitation in patients with chronic heart failure: A dutch practice guideline | Netherlands Heart Journal/ Dutch Royal Society for Physiotherapy (KNGF)/ Netherlands | Guideline | Chronic heart failure | Physiotherapist | Y | COI: N<br><br>FUND: Dutch Royal Society for Physiotherapy (KNGF) |
| Moe (2014) | The 2013 Canadian cardiovascular society heart failure management guidelines update: Focus on rehabilitation and exercise and surgical coronary revascularization | Canadian Journal of Cardiology / Canadian Cardiovascular Society - Heart Failure Management / Canada | Guidelines/ statement | Heart failure: frails, with preserved ejection fraction, with cardiac resynchronization therapy and implantable cardioverter defibrillators... | Clinicians and health care workers | N | COI: D<br><br>FUND: Canadian Cardiovascular Society |

| | | | | | | | |
|---|---|---|---|---|---|---|---|
| Achttien (2013) | Exercise-based cardiac rehabilitation in patients with coronary heart disease: A practice guideline | Netherlands Heart Journal / Dutch Royal Society for Physiotherapy (KNGF) / Netherlands | Guideline | Coronary heart disease | Physiotherapist | Y | COI: N<br><br>FUND: N |
| NICE (2013) | MI - Secondary Prevention: Secondary Prevention in Primary and Secondary Care for Patients Following a Myocardial Infarction: Partial Update of NICE CG48 | https://www.nice.org.uk / National Institute for Health and Care Excellence / England, UK | Guideline | Myocardial infarction | HCP | Y | COI: NR<br><br>FUND: NICE |
| Achttien (2011) | KNGF guideline: cardiac rehabilitation | Nederlands Tijdschrift voor Fysiotherapie [Dutch Journal of Physical Therapy] / Dutch Royal Society for Physiotherapy (KNGF) / Netherlands | Guideline | CHD patients [acute coronary syndrome: myocardial infarction, unstable angina pectoris, post percutaneous coronary intervention, post-CAPG or valve replacement] and Heart failure patients | Physiotherapist | Y | COI: NR<br><br>FUND: NR |
| Stone et al. (2009) | Canadian Guidelines for Cardiac Rehabilitation and Cardiovascular Disease Prevention | None / Canadian Association of Cardiac Rehabilitation / Canada | Guideline | Individuals with or without cardiovascular diseases, but is focused on CR patients | Healthcare practitioners working with cardiac rehabilitation | Y | COI: available from CACPR upon written request<br><br>Fund: available from CACPR upon written request |
| Hayman (2006) | Strengthening cardiac rehabilitation and secondary prevention for Aboriginal and Torres Strait Islander peoples | Medical Journal of Australia / Cardiac Rehabilitation Working Committee of the National Health and Rehabilitation Council / Australia | A Guide | Patients who have experienced cardiac events or procedures | Health professionals (in primary care, in hospitals), for managers | Y | COI: NR<br><br>FUND: NR |

| | | | | | | | |
|---|---|---|---|---|---|---|---|
| No author (2006) | Linee guida nazionali su cardiologia riabilitativa e prevenzione secondaria delle malattie cardiovascolari | Monaldi Arch Chest Dis / L'Agenzia per i Servizi Sanitari Regionali / Italy | Guideline | Patients with acute myocardial infarction (AMI) or a coronary revascularization, angina or heart failure and, post-AMI or post-angioplasty patients with complications, aortocoronary bypass or valve replacement surgery; patients with chronic heart failure, threatening arrhythmias and cardiac stimulator carriers (pacemakers, ventricular resynchronization, implantable defibrillators). | These guidelines are addressed for both citizens and to those who deal with CR (cardiologists, nurses, physiotherapists, physiatrists, dieticians, psychologists, occupational therapists) and to specialists and general practitioners (GPs) who place the indication. They can also be useful for those involved in health planning, costs and planning of interventions. | Y | COI: NR<br><br>FUND: Partially funded by PNLG, Istituto Superiore di Sanità |

COI: Conflict of interests; CR: cardiovascular rehabilitation; D: disclosed; HCP: healthcare professionals; N: nothing to declare; NR: not reported.

#### Rapid review of HIIT interventions

*Table S3. Quality assessment*

| Author, year | Population | Design | Q1 | Q2 | Q3 | Q4 | Q5 | Q6 | Q7 | Q8 | Q9 | Q10 | Q11 | Q12 | Total | Quality class |
|---|---|---|---|---|---|---|---|---|---|---|---|---|---|---|---|---|
| Boidin, 2019 | ACS | RCT | 1 | 1 | 0 | 1 | 1 | 3 | 0 | 2 | 1 | 0 | 0 | 1 | 11 | HIGHER |
| Aamot, 2014 | CAD | RCT | 1 | 1 | 1 | 0 | 0 | 3 | 1 | 2 | 1 | 1 | 1 | 1 | 13 | HIGHER |
| Conraads, 2015 | CAD | RCT | 1 | 0 | 1 | 0 | 1 | 2 | 1 | 2 | 1 | 0 | 0 | 1 | 10 | HIGHER |
| Currie, 2013 | CAD | Pre and post study | 1 | 0 | 1 | 1 | 0 | 3 | 0 | 2 | 1 | 0 | 0 | 1 | 10 | HIGHER |
| Currie, 2015 | CAD | RCT (factorial repeated-measures design) | 1 | 0 | 1 | 1 | 0 | 2 | 0 | 1 | 1 | 0 | 1 | 1 | 9 | HIGHER |
| Helgerud, 2011 | CAD | RCT | 1 | 0 | 1 | 1 | 0 | 2 | 0 | 2 | 1 | 0 | 1 | 1 | 10 | HIGHER |
| Karlsen, 2008 | CAD | RCT | 1 | 0 | 0 | 0 | 0 | 2 | 0 | 2 | 1 | 0 | 1 | 1 | 8 | HIGHER |
| Madssen, 2014 | CAD | RCT | 1 | 1 | 0 | 0 | 1 | 3 | 1 | 2 | 1 | 0 | 0 | 1 | 11 | HIGHER |
| Rognmo, 2004 | CAD | RCT | 1 | 1 | 1 | 1 | 1 | 3 | 0 | 1 | 1 | 0 | 0 | 1 | 11 | HIGHER |
| Szmigielska, 2018 | CAD | Pre post | 1 | 0 | 0 | 0 | 0 | 0 | 0 | 2 | 0 | 0 | 0 | 1 | 4 | LOWER |
| Taraldsen, 2020 | CAD | RCT | 1 | 1 | 1 | 0 | 0 | 2 | 0 | 2 | 1 | 0 | 0 | 1 | 9 | HIGHER |
| Villelabeitia-Jaureguizar, 2019 | CAD | RCT | 1 | 0 | 0 | 1 | 0 | 1 | 0 | 2 | 1 | 0 | 1 | 1 | 8 | HIGHER |
| Abdelhalem, 2018 | CAD | RCT | 1 | | 0 | 1 | 0 | 1+0+1 | 0 | 2 | 1 | 0 | 0 | 1 | 8 | HIGHER |
| Keteyian, 2014 | CAD | RCT | 1 | 1 | 0 | 0 | 1 | 1 | 0 | 2 | 1 | 0 | 1 | 1 | 9 | HIGHER |
| Lee, 2019 | CAD + menopause | RCT | 1 | 1 | 1 | 0 | 1 | 3 | 1 | 2 | 1 | 0 | 1 | 1 | 13 | HIGHER |
| Olsen, 2015 | CAD + obesity | RCT | 1 | 0 | 1 | 0 | 0 | 1 | 1 | 2 | 1 | 0 | 0 | 1 | 8 | HIGHER |
| Perdersen, 2015 CUT-IT trial | CAD + overweight | RCT | 1 | 0 | 1 | 0 | 0 | 2 | 1 | 2 | 1 | 0 | 0 | 1 | 9 | HIGHER |
| Cardozo, 2015 | CHD | RCT | 1 | 0 | 1 | 1 | 0 | 1 | 0 | 2 | 1 | 0 | 0 | 1 | 8 | HIGHER |

| Author, year | Population | Design | Q1 | Q2 | Q3 | Q4 | Q5 | Q6 | Q7 | Q8 | Q9 | Q10 | Q11 | Q12 | Total | Quality class |
|---|---|---|---|---|---|---|---|---|---|---|---|---|---|---|---|---|
| Dobraszkiewicz-Wasilewska, 2009 | CHD | Pre and post study | 1 | 0 | 0 | 0 | 0 | 0 | 0 | 0 | 1 | 0 | 0 | 1 | 3 | LOWER |
| Rognmo, 2012 | CHD | Retrospective - database | 0 | 0 | 0 | 0 | 0 | 1 | 0 | 1 | 0 | 0 | 0 | 1 | 3 | LOWER |
| Moholdt, 2012 | MI | RCT | 1 | 1 | 0 | 0 | 0 | 2 | 1 | 2 | 1 | 0 | 0 | 1 | 9 | HIGHER |
| Santi, 2018 | MI | RCT | 1 | 0 | 0 | 0 | 0 | 0 | 0 | 2 | 1 | 0 | 0 | 1 | 5 | LOWER |
| Dun, 2019 | MI + metabolic syndrome | Retrospective - parallel groups | 1 | 0 | 0 | 1 | 0 | 3 | 0 | 2 | 1 | 0 | 1 | 1 | 10 | HIGHER |
| Choi, 2018 | MI + PCI | RCT | 1 | 0 | 1 | 1 | 0 | 3 | 0 | 2 | 1 | 0 | 0 | 1 | 10 | HIGHER |
| Munk, 2009 | PCI | RCT | 1 | 1 | 1 | 1 | 0 | 3 | 0 | 2 | 1 | 0 | 0 | 1 | 11 | HIGHER |
| Malmo, 2016 | AF | RCT | 1 | 0 | 0 | 0 | 1 | 0+1+1 | 0 | 1+1 | 1 | 1 | 0 | 1 | 9 | HIGHER |
| Skielboe, 2017 | AF | RCT | 1 | 1 | 1 | 1 | 1 | 1+1+0 | 1 | 1+1 | 1 | 0 | 1 | 1 | 13 | HIGHER |
| Roditis, 2007 | CHF | RCT | 0 | 0 | 0 | 0 | 0 | 1+0+1 | 0 | 1+1 | 1 | 0 | 1 | 1 | 8 | HIGHER |
| Spee, 2016 | CHF | RCT | 1 | 1 | 1 | 1 | 1 | 1+1+1 | 0 | 1+1 | 1 | 0 | 0 | 1 | 12 | HIGHER |
| Novaković, 2018 | ConHD | RCT | 1 | 1 | 1 | 1 | 0 | 1+1+1 | 0 | 1+1 | 1 | 1 | 1 | 1 | 13 | HIGHER |
| Sandberg, 2018 | ConHD | RCT | 1 | 1 | 1 | 1 | 0 | 1+1+1 | 0 | 1+1 | 1 | 0 | 1 | 1 | 12 | HIGHER |
| Anagnostakou, 2011 | HF | RCT | 1 | 1 | 0 | 1 | 1 | 0+1+0 | 0 | 1+1 | 1 | 0 | 1 | 1 | 10 | HIGHER |
| Chou, 2019 | HF | RCT | 1 | 1 | 0 | 1 | 1 | 1+0+0 | 0 | 1+1 | 1 | 1 | 1 | 1 | 11 | HIGHER |
| Chrysohoou, 2014 | HF | RCT | 1 | 1 | 1 | 0 | 0 | 0+1+1 | 1 | 1+1 | 1 | 0 | 0 | 1 | 10 | HIGHER |
| Dimopoulos, 2006 | HF | RCT | 1 | 1 | 0 | 1 | 0 | 0+0+1 | 0 | 1+1 | 1 | 0 | 1 | 1 | 9 | HIGHER |

| Author, year | Population | Design | Q1 | Q2 | Q3 | Q4 | Q5 | Q6 | Q7 | Q8 | Q9 | Q10 | Q11 | Q12 | Total | Quality class |
|---|---|---|---|---|---|---|---|---|---|---|---|---|---|---|---|---|
| Ellingsen, 2017 | HF | RCT | 1 | 0 | 0 | 0 | 1 | 0+1+1 | 1 | 1+1 | 1 | 0 | 0 | 1 | 9 | HIGHER |
| Hsu, 2019 | HF | prospective pre and post study (matched-control study) | 1 | 0/NA | 1 | 1 | 0 | 1+0+0 | 0 | 1+1 | 1 | 0 | 0 | 1 | 8 | HIGHER |
| Huang, 2014 | HF | pre and post study | 1 | 0 | 0 | 1 | 0 | 0+0+1 | 0 | 1+1 | 1 | 0 | 0 | 1 | 7 | LOWER |
| Iellamo, 2014 | HF | RCT | 1 | 1 | 0 | 1 | 0 | 1+1+1 | 0 | 1+1 | 1 | 0 | 0 | 0 | 9 | HIGHER |
| Koufaki, 2014 | HF | RCT | 1 | 0 | 0 | 1 | 1 | 0+0+1 | 0 | 1+1 | 1 | 0 | 0 | 1 | 8 | HIGHER |
| Sadek, 2020 | HF | RCT | 1 | 1 | 0 | 1 | 1 | 1+1+0 | 0 | 1+1 | 1 | 1 | 0 | 1 | 12 | HIGHER |
| Thijssen, 2019 | HF | Non-randomized clinical trial | 1 | 0 | 1 | 1 | 1 | 1+0+1 | 1 | 1+1 | 1 | 0 | 1 | 1 | 12 | HIGHER |
| Tzanis, 2017 | HF | Randomized parallel 2-group study | 0 | 0 | 0 | 0 | 0 | 1+1+1 | 0 | 1+1 | 1 | 0 | 0 | 1 | 7 | LOWER |
| Wisløff, 2007 | HF | RCT | 1 | 1 | 1 | 1 | 1 | 1+1+1 | 0 | 1+1 | 1 | 0 | 1 | 1 | 13 | HIGHER |
| Melo, 2019 | HF + AF | RCT | 1 | 1 | 0 | 0 | 0 | 0+0+1 | 0 | 1+1 | 1 | 0 | 0 | 1 | 7 | LOWER |
| Santa-Clara, 2019 | HF + CRT | RCT | 0 | 1 | 1 | 1 | 0 | 1+1+1 | 1 | 1+1 | 1 | 0 | 0 | 1 | 11 | HIGHER |
| Spee, 2020 | HF + CRT | RCT | 1 | 1 | 1 | 1 | 1 | 1+1+1 | 0 | 1+1 | 1 | 0 | 0 | 1 | 12 | HIGHER |
| Isaksen,2015 | HF + ICD or CRT-D | pre and post study | 1 | 0 | 0 | 1 | 1 | 1+1+1 | 0 | 1+1 | 1 | 0 | 1 | 1 | 11 | HIGHER |
| Moreno-Suarez, 2020 | HF + LVAD | RCT | 1 | 1 | 1 | 1 | 1 | 0+1+1 | 1 | 1+1 | 1 | 0 | 1 | 1 | 13 | HIGHER |
| Fu, 2013 | HF | RCT | 1 | 0 | 1 | 0 | 0 | 1+0+0 | 0 | 1+1 | 1 | 0 | 1 | 1 | 8 | HIGHER |

| Author, year | Population | Design | Q1 | Q2 | Q3 | Q4 | Q5 | Q6 | Q7 | Q8 | Q9 | Q10 | Q11 | Q12 | Total | Quality class |
|---|---|---|---|---|---|---|---|---|---|---|---|---|---|---|---|---|
| Fu, 2016 | HF | pre and post study | 1 | 0 | 0 | 1 | 1 | 1+0+0 | 0 | 1+1 | 1 | 0 | 1 | 1 | 9 | HIGHER |
| Donelli da Silveira, 2020 | HFpEF | RCT | 1 | 1 | 1 | 1 | 1 | 0+1+1 | 0 | 1+1 | 1 | 0 | 1 | 1 | 12 | HIGHER |
| Hsu, 2020 | HFrEF | pre and post study | 1 | 0/NA | 0 | 0 | 1 | 0+1+0 | 0 | 1+1 | 1 | 0 | 1 | 1 | 8 | HIGHER |
| Dall, 2015 | HTx | RCT | 1 | 1 | 1 | 0 | 0 | 1 | 0 | 0+0 | 1 | 0 | 0 | 1 | 6 | LOWER |
| Nytrøen, 2012 | HTx | RCT | 1 | 1 | 1 | 1 | 0 | 1+1+1 | 0 | 1+1 | 1 | 0 | 1 | 1 | 11 | HIGHER |
| Way, 2020 | CVD | Retrospective mixed-methods analysis | 1 | 0/NA-retrospective study | 0/NA-retrospective study | 0/NA-retrospective study | 0/NA-retrospective study | 1+1+1 | 0 | 0 | 1 | 0 | 1 | 1 | 7 | LOWER |
| Alshamari, 2023 | HF | RCT | 1 | 1 | 1 | 1 | 0 | 0+1+0 | 0 | 1+1 | 1 | 0 | 1 | 1 | 10 | HIGHER |
| Avazpour, 2023 | CHD | Clinical trial with a pre-test-post-test | 1 | 0 | 1 | 1 | 0 | 0+0+0 | 0 | 1+1 | 1 | 0 | 1 | 1 | 8 | HIGHER |
| Dun, 2021 | MI | Prospective cohort | 1 | 0 | 0 | 0 | 0 | 0+1+0 | 0 | 0/NA | 1 | 0/NA | 1 | 1 | 5 | LOWER |
| Dunford, 2021 | MI | RCT | 1 | 1 | 1 | 1 | 0 | 1+1+1 | 0 | 1+1 | 1 | 0 | 1 | 1 | 12 | HIGHER |
| Hsu, 2023 | HF | Prospective cohort | 1 | 0 | 1 | 0/NA | 0 | 0+0+1 | 0 | 0/NA | 1 | NA | 0 | 1 | 5 | LOWER |
| Kim, 2023 | Symptomatic non-permanent AF | RCT | 1 | 1 | 1 | 1 | 1 | 1+1+1 | 1 | 1+1 | 1 | 0 | 1 | 1 | 14 | HIGHER |
| Koppen, 2021 | HFrEF | RCT | 1 | 1 | 1 | 1 | 1 | 0+1+1 | 1 | 1+0 | 1 | 0 | 0 | 1 | 11 | HIGHER |
| Kristiansen, 2022 | CAD | RCT | 1 | 1 | 1 | 1 | 0 | 1+1+1 | 1 | 1+1 | 1 | 0 | 1 | 1 | 13 | HIGHER |

| Author, year | Population | Design | Q1 | Q2 | Q3 | Q4 | Q5 | Q6 | Q7 | Q8 | Q9 | Q10 | Q11 | Q12 | Total | Quality class |
|---|---|---|---|---|---|---|---|---|---|---|---|---|---|---|---|---|
| Larsen, 2023 | ANOCA | RCT | 1 | 0 | 0 | 0 | 0 | 1+1+1 | 0 | 1+1 | 1 | 0 | 0 | 1 | 8 | HIGHER |
| Lund, 2020 | Acute MI | single group pre-post interventional study | 0 | 0 | 0 | NA | 1 | 0+1+1 | 0 | 0/NA | 1 | 0/NA | 0 | 1 | 5 | LOWER |
| Lundgren, 2023 | HF | RCT | 1 | 0 | 1 | 1 | 1 | 1+1+1 | 0 | 1+1 | 1 | 0 | 0 | 1 | 11 | HIGHER |
| McGregor, 2023 | MI, CABG, CAD, PCI | RCT | 1 | 1 | 1 | 1 | 1 | 0+1+1 | 1 | 1+1 | 0 | 0 | 1 | 1 | 12 | HIGHER |
| Mueller, 2021 | HFpEF | RCT | 0 | 1 | 1 | 0 | 1 | 1+1+0 | 0 | 1+1 | 1 | 0 | 1 | 1 | 10 | HIGHER |
| Reed (1), 2022 | CAD | RCT | 1 | 1 | 1 | 1 | 1 | 0+1+1 | 1 | 1+1 | 1 | 0 | 0 | 1 | 12 | HIGHER |
| Reed (2), 2022 | AF | RCT | 1 | 1 | 1 | 1 | 1 | 0+0+1 | 1 | 1 | 1 | 1 | 1 | 1 | 12 | HIGHER |
| Rodrigo Aispuru-Lanche, 2023 | MI | RCT | 1 | 0 | 1 | 1 | 1 | 0+1+1 | 1 | 1+1 | 1 | 0 | 0 | 1 | 11 | HIGHER |
| Sadek, 2023 | HF+ IMW | RCT | 1 | 0 | 0 | 1 | 1 | 1+1+1 | 1 | 1 | 1 | 0 | 0 | 1 | 10 | HIGHER |
| Sales, 2020 | HFrEF | RCT | 1 | 0 | 0 | 1 | 0 | 1+0+0 | 0 | 1 | 1 | 0 | 1 | 1 | 7 | LOWER |
| Szmigielska, 2022 | ACS | Prospective cohort | 0 | 0 | NA | 0/NA | 0 | 0+0+0 | 0 | 1+1 | 1 | 0 | 1 | 1 | 5 | LOWER |
| Taylor, 2020 | CAD | RCT | 1 | 0 | 1 | 1 | 0 | 1+1+1 | 1 | 1+1 | 1 | 0 | 1 | 1 | 12 | HIGHER |
| Turri-Silva, 2021 | HF | RCT | 1 | 1 | 1 | 1 | 1 | 1+1+1 | 0 | 1+1 | 1 | 0 | 0 | 1 | 12 | HIGHER |
| Vesterbekkmo, 2023 | CAD | RCT | 1 | 1 | 1 | 1 | 1 | 0+1+0 | 1 | 1+1 | 1 | 0 | 0 | 1 | 11 | HIGHER |
| Vidal-Almela, 2022 | CVD | Prospective pre-post study | 1 | NA | NA | NA | 0 | NA+0+1 | NA | NA | 1 | NA | 0 | 1 | 4 | LOWER |

| Author, year | Population | Design | Q1 | Q2 | Q3 | Q4 | Q5 | Q6 | Q7 | Q8 | Q9 | Q10 | Q11 | Q12 | Total | Quality class |
|---|---|---|---|---|---|---|---|---|---|---|---|---|---|---|---|---|
| Winzer, 2022 | HFpEF | RCT | 1 | 0 | 1 | 1 | 1 | 0+0+0 | 0 | 1+1 | 1 | 0 | 1 | 1 | 9 | HIGHER |
| Yakut, 2022 | MI | RCT | 1 | 1 | 1 | 1 | 0 | 1+1+0 | 0 | 1+1 | 1 | 0 | 1 | 1 | 11 | HIGHER |

Q1: Eligibility criteria specified Eligibility criteria should be specified and fulfilled and specific diagnostic test values should be provided for all participants.
Q2: Randomization specified A description of the method used to allocate patients into treatment groups should be provided.
Q3: Allocation concealment It should be stated if group allocation was concealed; meaning if a patient was eligible for inclusion in the trial was unaware (when this decision was made) of which group the patient would be allocated to.
Q4: Groups similar at baseline data of all participants who were randomized should be presented. There should be no significant difference in the measure of the severity of the treated condition between treatment groups.
Q5: Blinding of assessor (for at least one key outcome)
Q6: Outcome measures assessed in 85% of patients
Q7: Intention-to-treat analysis When a patient withdraws, this analysis is conducted by using either the last value obtained for each of the outcome measures as a post-intervention value, or by using the baseline value as a post value.
Q8: Between-group statistical comparisons reported
Q9: Point measures and measures of variability for all reported outcome measures
Q10: Activity monitoring in control groups
Q11: Relative exercise intensity remained constant
Q12: Exercise volume and energy expenditure
**ACS**: acute coronary syndrome; **AF**: atrial fibrillation; **ANOCA**: angina with no obstructive coronary artery disease; **CABG**: coronary artery bypass graft; **CAD**: coronary artery disease; **CHD**: congenital heart disease; **ConHD**: congenital heart disease; **CRT**: Cardiac resynchronization therapy; **CRT-D**: Cardiac resynchronization therapy with a defibrillator; **CVD**: cardiovascular disease; **HF**: heart failure; **HFpEF**: heart failure with preserved ejection fraction; **HFrEF**: heart failure with reduced ejection fraction; **HTx**: heart transplant; **IMW**: inspiratory muscle weakness; **LVAD**: Cardiac resynchronization therapy with a defibrillator; **MI**: myocardial infarction; **PCI**: percutaneous coronary intervention; **RCT**: randomised control trial

*Table S4. List of studies by patient profile (N=81)*

| Author, year | Population | Sample size, sex | Study design | Number of arms | Intervention duration | Intervention site | Target intensity | HIIT Prescription | Training volume (HIIT group) | Ergometer - Exercise testing | Exercise capacity / Functional assessment tool | Ergometer - HIIT |
|---|---|---|---|---|---|---|---|---|---|---|---|---|
| Boidin, 2019 | ACS | HIIT short: 24 (13 male, 5 female); MICT: 19 (16 male, 3 female) | RCT | 2: HIIT short vs MICT | 12 weeks | Centre | 100% PPO | 3 sets of 10 min of repeated phases of 15 seconds at 100% of peak power output alternating with 15 seconds of passive recovery (complete rest). The three sets were separated by 4 minutes of passive recovery. | 1368 min | Cycle ergometer | CPET | Bicycle ergometer |
| Malmo, 2016 | AF | HIIT long: 26 (20 male, 77%); CG: 25 (22 male, 88%) | RCT | 2: HIIT long vs CG | 12 weeks | Centre + home | 85% to 95% of HRpeak | 4 times 4 min intervals with 3 minutes of active recovery | 1008 min | Treadmill | CPET | Treadmill |

| Author, year | Population | Sample size, sex | Study design | Number of arms | Intervention duration | Intervention site | Target intensity | HIIT Prescription | Training volume (HIIT group) | Ergometer - Exercise testing | Exercise capacity / Functional assessment tool | Ergometer - HIIT |
|---|---|---|---|---|---|---|---|---|---|---|---|---|
| Skielboe, 2017 | AF | HIIT (*interval length NR*): 37 (22 male, 59.4%); Low intensity 33 (19 male, 57.6%) | RCT | 2: HIIT vs low intensity | 12 weeks | Hospital | 80% of maximal perceived exertion | 20 min interval exercising on ergometer bike (with allocated intensity), 20 min varying circuit exercise on the floor (with allocated intensity) | 960 min | Cycle ergometer | CPET | Cycle ergometer |
| Aamot, 2014 | CAD | HIIT long in group: 28 (25 male, 3 female); HIIT long treadmill: 34 (28 male, 6 female); HIIT long HOME: 28 (27 male, 1 female) | RCT | 3: HIIT long in Group vs Treadmill vs Home | 12 weeks | Home-based vs hospital | 85-95% HRpeak | 4 intervals lasting 4 minutes each, separ ated by 4 minutes of active breaks | 768 min | Treadmill | CPET | Treadmill, cross trainer |

| Author, year | Population | Sample size, sex | Study design | Number of arms | Intervention duration | Intervention site | Target intensity | HIIT Prescription | Training volume (HIIT group) | Ergometer - Exercise testing | Exercise capacity / Functional assessment tool | Ergometer - HIIT |
|---|---|---|---|---|---|---|---|---|---|---|---|---|
| Conraads, 2015 | CAD | HIIT long: 100 (91 male, 9 female); MICT: 100 (89 male, 11 female) | RCT | 2: HIIT long vs MICT | 12 weeks | Centre | 90-95% HRpeak | 4 min interval with 3 min interval of active breaks | 1008 min | Cycle ergometer | CPET | Bicycle |
| Currie, 2013 | CAD | HIIT short: 11; MICT: 11 (total 24 male, 2 female) | Pre and post study | 2: HIIT short vs MICT | 12 weeks | Centre | 89% PPO (range, 80%–104%) | Active and break intervals of 1 min | 480 min | Cycle ergometer | CPET | Cycle ergometer |

| **Author, year** | **Population** | **Sample size, sex** | **Study design** | **Number of arms** | **Intervention duration** | **Intervention site** | **Target intensity** | **HIIT Prescription** | **Training volume (HIIT group)** | **Ergometer - Exercise testing** | **Exercise capacity / Functional assessment tool** | **Ergometer - HIIT** |
|---|---|---|---|---|---|---|---|---|---|---|---|---|
| Currie, 2015 | CAD | HIIT short: 9; MICT: 10 (total 26 male, 1 female) | RCT (factorial repeated - measures design) | 2: HIIT short vs MICT | 12 weeks | Centre | 85% PPO (75-93%), 100% PPOpre for month 2 and 108% PPOpre for month 3. During the final 3 months, the HIIT group trained at 121% (100–152%) of PPOpre | Active and break intervals of 1 min | 960 min | Cycle ergometer | CPET | Cycle ergometer |
| Helgerud, 2011 | CAD | HIIT long: 8 (6 male, 2 female); Strength training: 10 (10 make) | RCT | 2: HIIT long vs maximal strength training group (MST) | 8 weeks | *Unclear, hospital* | 85-95 % HRpeak | 4 times 4 min of interval training, with 3 min active breaks | 840 min | Treadmill | CPET | Treadmill |

| Author, year | Population | Sample size, sex | Study design | Number of arms | Intervention duration | Intervention site | Target intensity | HIIT Prescription | Training volume (HIIT group) | Ergometer - Exercise testing | Exercise capacity / Functional assessment tool | Ergometer - HIIT |
|---|---|---|---|---|---|---|---|---|---|---|---|---|
| Karlsen, 2008 | CAD | HIIT long normoxic: 10 (7 male, 3 female); HIIT long hyperoxic: 8 (6 male, 2 female) | RCT | 2: HIIT long hyperoxic and normoxic | 10 weeks | Centre | 85-95% HRpeak | 4 intervals of 4 min, with 3 min active breaks | 840 min | Treadmill | CPET | Treadmill |
| Madssen, 2014 | CAD | HIIT long: 15 (14 male, 1 female); MICT: 21 (15 male, 6 female) | RCT | 2: HIIT long vs MICT | 12 weeks | Centre | 85-95% HRpeak | 4 intervals of 4 min, with 3 min active breaks | 1008 min | Treadmill | CPET | Treadmill |
| Rognmo, 2004 | CAD | HIIT long: 8 (6 male, 2 female); MICT: 9 (8 male, 1 female) | RCT | 2: HIIT long vs MICT | 10 weeks | centre | 80-90% $\dot{V}O_2max$ / 85-95% HR peak | 4 intervals of 4 min, with 3 min active breaks | 840 min | Treadmill | CPET | Treadmill |
| Szmigielska, 2018 | CAD | HIIT long: 131 (all male) | Pre post | 1: HIIT long | 8 weeks | Centre | 14 - 16 RPE | 4 min of workload and 2 min of active breaks | 1080 min | Cycle ergometer | CPET | Cycle ergometer |

| Author, year | Population | Sample size, sex | Study design | Number of arms | Intervention duration | Intervention site | Target intensity | HIIT Prescription | Training volume (HIIT group) | Ergometer - Exercise testing | Exercise capacity / Functional assessment tool | Ergometer - HIIT |
|---|---|---|---|---|---|---|---|---|---|---|---|---|
| Taraldsen, 2020 | CAD | HIIT long: 14; MICT: 18 | RCT | 2: HIIT long vs MICT | 12 weeks | Centre | 85-95% HRpeak | Four times 4-min high intensity intervals with 3 min of active rest between each interval and before terminating the session. | 1008 min | Treadmill | CPET | NR |
| Villelabeitia-Jaureguizar, 2019 | CAD | HIIT short: 57 (50, 87.7% male); MICT: 53 (42, 79.2% male) | RCT | 2: HIIT short vs MICT + walking in unsupervised days | 8 weeks | Centre | 50% of the maximum load reached with the SRT (peak intervals) were followed by recovery periods at 10% | 20-s of active period intercalated with 40-s recovery periods | 720 min | Cycle ergometer | CPET | Cycle ergometer |

| Author, year | Population | Sample size, sex | Study design | Number of arms | Intervention duration | Intervention site | Target intensity | HIIT Prescription | Training volume (HIIT group) | Ergometer - Exercise testing | Exercise capacity / Functional assessment tool | Ergometer - HIIT |
|---|---|---|---|---|---|---|---|---|---|---|---|---|
| Abdelhalem, 2018 | CAD | HIIT progressive: 20 (18, 90% male; 2, 10% female); MICT: 20 (16, 80% male; 4, 20% female) | RCT | 2: HIIT progressive vs MICT | 12 weeks | Centre | 85-95% HRR | Alternating brief 2–5 min | 840 min | Treadmill | GXT | NR |
| Keteyian, 2014 | CAD | HIIT long: 21 (11, 73% male); MICT: 18 (12, 92% male) | RCT | 2: HIIT long vs MICT | 10 weeks | Centre | 80-90% HRR | 4 intervals of 4 min, with 3 min active breaks | 840 min | Treadmill | CPET | Treadmill |
| Lee, 2019 | CAD + menopause | HIIT long: 17; MICT: 14 (31, 100% female) | RCT | 2: HIIT long vs MICT | 24 weeks | Mixed: Centre and home | 60–80% $\dot{V}O_2$peak | 4 intervals of 4 min, with 3 min active breaks | 840 min | Cycle ergometer | CPET | Track or treadmill |

| Author, year | Population | Sample size, sex | Study design | Number of arms | Intervention duration | Intervention site | Target intensity | HIIT Prescription | Training volume (HIIT group) | Ergometer - Exercise testing | Exercise capacity / Functional assessment tool | Ergometer - HIIT |
|---|---|---|---|---|---|---|---|---|---|---|---|---|
| Olsen, 2015 | CAD + obesity | HIIT (*interval length NR*): 26 (22, 85% male); LED: 29 (21, 72% male) | RCT | 2: HIIT vs LED (diet) | 12 | centre | 85-90% V̇O₂peak | 1–4 min intervals, with a total of 16 min, separated by active pauses of 1–3 min | 576 min | Cycle ergometer | CPET | NR |
| Perdersen, 2015 | CAD + overweight | HIIT mixed: 35 (22, 85% male); LED: 35 (21, 72% male) | RCT | 2: HIIT mixed vs LED (diet) | 12 | centre | 90% HR peak abstract | 1–4 min intervals separated by active pauses of 1–3 min | NR | Cycle ergometer | CPET | NR |
| Cardozo, 2015 | CHD | HIIT medium: 23 (63% male); MICT: 24 (66% male); CG: 24 (76% male) | RCT | 3: HIIT medium* vs MICT vs CG | 16 weeks | Centre | 90% HRpeak | Active and recovery periods alternated every 2 min | 1440 min | Treadmill | CPET | Treadmill |

| Author, year | Population | Sample size, sex | Study design | Number of arms | Intervention duration | Intervention site | Target intensity | HIIT Prescription | Training volume (HIIT group) | Ergometer - Exercise testing | Exercise capacity / Functional assessment tool | Ergometer - HIIT |
|---|---|---|---|---|---|---|---|---|---|---|---|---|
| Dobraszkiewicz-Wasilewska, 2009 | CHD | HIIT long: 151 males after MI (90 noninvasive therapy; 61 CABG) | Pre and post study | 2: HIIT long(with the same intervention) (subgroup after a MI subjected to, noninvasive therapy; people treated by means of a CABG) | 8 weeks | Centre | Training HR= 80% x (HRmax – HRrest) + HRrest) | 3 intervals of 4-min with 2 min resting interval | 288 min | Treadmill | CPET | Cycle ergometer |

| **Author, year** | **Population** | **Sample size, sex** | **Study design** | **Number of arms** | **Intervention duration** | **Intervention site** | **Target intensity** | **HIIT Prescription** | **Training volume (HIIT group)** | **Ergometer - Exercise testing** | **Exercise capacity / Functional assessment tool** | **Ergometer - HIIT** |
|---|---|---|---|---|---|---|---|---|---|---|---|---|
| Rognmo, 2012 | CHD (myocardial infarction, angioplasty, coronary surgery, valve surgery, heart failure) | N = 4846 (a total of 175 820 exercise sessions lasting approximately one hour were recorded, distributed on 129 456 hours of moderate-intensity exercise and 46 364 hours of high-intensity exercise, respectively.)<br><br>The sample was 70% male and 30% female) | Retrospective - database | 2: HIIT long vs MICT | NR | Centre | 85-95% HRpeak | Four 4-minute high intensity intervals. Each interval was separated by active pauses | NR | NR | CPET, only included for safety | Typically consisted of treadmill exercise, but aerobic group training, biking sessions, and outdoor walking and cross-country skiing were also performed. |

| Author, year | Population | Sample size, sex | Study design | Number of arms | Intervention duration | Intervention site | Target intensity | HIIT Prescription | Training volume (HIIT group) | Ergometer - Exercise testing | Exercise capacity / Functional assessment tool | Ergometer - HIIT |
|---|---|---|---|---|---|---|---|---|---|---|---|---|
| Novaković, 2018 | ConHD | HIIT short: 9; MICT: 9; CG: 9 Total 17, 63% female | RCT | 3: HIIT short vs MICT vs CG | 12-18 weeks | Centre | 80% HRpeak | 1-minute exercise followed by 3-minute exercise recovery | 1152 min | Cycle ergometer | CPET | Cycling and/or speed walking |
| Sandberg, 2018 | ConHD | HIIT progressive: 13 (8, 62%) male); CG: 10 (4, 40% male) | RCT | 2: HIIT progressive vs CG | 12 weeks | Home-based | 75%-80% HRmax (Karvonen) - Borg 15-16 | Interval duration was individually adjusted. The maximum interval time was 5 min, with active recovery periods of 3 min | NR | NR | CPET | Cycle ergometer |
| Dimopoulos, 2006 | HF | HIIT short: 10 (9 male, 90%); MICT: 14 (100% male) | RCT | 2: HIIT short vs MICT | 12 weeks | Centre | 100% baseline WRp | 30 s of exercise with 30 s of rest | 1440 min | Cycle ergometer | CPET | Cycle ergometer |

| Author, year | Population | Sample size, sex | Study design | Number of arms | Intervention duration | Intervention site | Target intensity | HIIT Prescription | Training volume (HIIT group) | Ergometer - Exercise testing | Exercise capacity / Functional assessment tool | Ergometer - HIIT |
|---|---|---|---|---|---|---|---|---|---|---|---|---|
| Roditis, 2007 | HF | HIIT short: 11 (10 male, 1 female); MICT: 10 (9 male, 1 female) | RCT | 2: HIIT short vs MICT | 12 weeks | Centre | 100% of baseline pWR | 30 s of exercise with 30 s of rest | 1440 min | Cycle ergometer | CPET | Cycle ergometer |
| Fu, 2013 | HF (HFpEF and HFrEF) | HIIT long: 14 (9 male, 5 female); MICT: 13 (8 male, 5 female); GHC: 13 (9 male, 4 female) | RCT | 3: HIIT long, MICT, GHC | 12 weeks | Hospital | 80% $\dot{V}O_2$peak (≈80% HRR) | Five 3-min intervals separated by 3 min exercise at lower intensity | 1080 min | Cycle ergometer | CPET | Cycle ergometer |
| Chou, 2019 | HF (HFrEF or HFpEF) | HIIT long: 15 (11 male, 4 female); CG: 15 (11 male, 4 female) | RCT | 2: HIIT long vs CG | 12 weeks | Centre | 80% $\dot{V}O_2$peak | Five 3-min intervals separated by 3 min exercise at lower intensity | 1080 min | Cycle ergometer | CPET | Cycle ergometer |

| **Author, year** | **Population** | **Sample size, sex** | **Study design** | **Number of arms** | **Intervention duration** | **Intervention site** | **Target intensity** | **HIIT Prescription** | **Training volume (HIIT group)** | **Ergometer - Exercise testing** | **Exercise capacity / Functional assessment tool** | **Ergometer - HIIT** |
|---|---|---|---|---|---|---|---|---|---|---|---|---|
| Fu, 2016 | HF (HFrEF or HFpEF) | HIIT long HFpEF: 30 (10 male, 10 female); HIIT long HFrEF: 30 (21 male, 9 female); GHC HFpEF: 29 (18 male, 11 female); GHC HFrEF: 28 (18 male, 10 female) | RCT | 4: HFpEF HIIT long, HFrEF HIIT long, HFpEF GHC, HFrEF GHC | 12 weeks | Centre | 80% $\dot{V}O_2$peak (80% HRreserve) | Five 3-min intervals separated by 3-min active break | 1080 min | Cycle ergometer | CPET | Cycle ergometer |
| Hsu, 2019 | HF (HFrEF or HFpEF) | HIIT long + MDP: 101 (70 male, 31 female ); MDP: 101 (74 male, 27 female) | pre- and post-study | 2: HIIT long vs MDP (disease management program) | ±12 weeks (36 sessions, 2-3x/week ) | Centre | 80% $\dot{V}O_2$peak | Five 3-min intervals separated by 3-min active break | 1080 min | Cycle ergometer | CPET | Bicycle ergometer |

| Author, year | Population | Sample size, sex | Study design | Number of arms | Intervention duration | Intervention site | Target intensity | HIIT Prescription | Training volume (HIIT group) | Ergometer - Exercise testing | Exercise capacity / Functional assessment tool | Ergometer - HIIT |
|---|---|---|---|---|---|---|---|---|---|---|---|---|
| Santa-Clara, 2019 | HF + CRT | HIIT progressive: 34; Control: 29 | RCT | 2: HIIT progressive vs control | 24 weeks | Hospital | 90–95% peak HR | 4 interval training periods with 3 lower-intensity active periods. During the first month, each interval training and active pause was increased by 30 s on a weekly basis, until accomplishing the 4 min work with 3 min of active rest. | Approximately 1200 min (when considering the target interval duration) | Treadmill | CPET, only included for safety and telemetry | NR |
| Isaksen, 2015 | HF + ICD or CRT-D | HIIT long: 26 (88% male); CG: 12 (100% male) | pre- and post-study | 2: HIIT long vs CG | 12 weeks | Centre | 85% of HRmax | 4 intervals of 4 min, with 3 min active breaks | 1008 min | Cycle ergometer | CPET | Treadmill or cycle ergometer |

| **Author, year** | **Population** | **Sample size, sex** | **Study design** | **Number of arms** | **Intervention duration** | **Intervention site** | **Target intensity** | **HIIT Prescription** | **Training volume (HIIT group)** | **Ergometer - Exercise testing** | **Exercise capacity / Functional assessment tool** | **Ergometer - HIIT** |
|---|---|---|---|---|---|---|---|---|---|---|---|---|
| Moreno-Suarez, 2020 | HF + LVAD | HIIT long: 11 (27% female); MICT: 10 (50% female) | RCT | 2: HIIT long vs MICT | 12 weeks | Centre | 80% −90% $\dot{V}O_2$ reserve | 4 intervals of 4 min, with 3 min active breaks | 1008 min | Treadmill | CPET, 6MWT | Treadmill |
| Donelli da Silveira, 2020 | HFpEF | HIIT long: 12 (70% female); MICT: 12 (56% female) | RCT | 2: HIIT long vs MICT | 12 weeks | Centre | 80-90% VO2peak and 85-95% HRpeak, aiming RPE 15-17 | 4 intervals of 4 min, with 3 min active breaks | 900 min | Treadmill | CPET | Treadmill |
| Anagnostakou, 2011 | HFrEF | HIIT short: 14 (12 male, 2 female); HIIT + Strength: 14 (11 male, 3 female) | RCT | 2:HIIT short vs HIIT plus strength | 12 weeks | Centre | 50% workload SRT | 30 s of exercise - 60 s of complete rest | 1440 min | Cycle ergometer | CPET | Cycle ergometer |
| Chrysohoou, 2014 | HFrEF | HIIT short: 50 (88% male); CG: 50 (72% male) | RCT | 2: HIIT short vs CG | 12 weeks | *Unclear, hospital* | 80-100% WRpeak | 30 s of exercise - 30 s of rest | 1620 min | Cycle ergometer | CPET | NR |

| Author, year | Population | Sample size, sex | Study design | Number of arms | Intervention duration | Intervention site | Target intensity | HIIT Prescription | Training volume (HIIT group) | Ergometer - Exercise testing | Exercise capacity / Functional assessment tool | Ergometer - HIIT |
|---|---|---|---|---|---|---|---|---|---|---|---|---|
| Ellingsen, 2017 | HFrEF | HIIT long: 88 (18% female); MICT: 78 (19% female); CG: 81 (19% female) | RCT | 3: HIIT long , MICT, vs recommendation of regular exercise (RRE) | 12 weeks | Centre | 90-95% HRmax | 4 intervals of 4 min, with 3 min active breaks | 1008 min | Treadmill or Cycle ergometer | CPET | Treadmill or bicycle |
| Hsu, 2020 | HFrEF | HIIT long: 38 (22 male, 16 female); NC (22 male, 16 female) | pre and post study | 2: HIIT long vs NC (healthy control - did not receive any form of intervention) | 12 weeks | Centre | 80% $\dot{V}O_2$peak (≈80% HRR) | Five 3 min intervals separated by 3 min exercise at lower intensity | 1080 min | Cycle ergometer | CPET | Cycle ergometer |
| Huang, 2014 | HFrEF | mHIIT long: 33 (26 male, 7 female); UC: 33 (25 male, 8 female) | pre and post study | 2: mHIIT long vs UC (optimal medical treatment only) | 12 weeks | Centre | 80% $\dot{V}O_2$ reserve | Seven 3 min intervals separated by 3 min exercise at lower intensity | 1008 min | Cycle ergometer | CPET | Cycle ergometer or treadmill |

| Author, year | Population | Sample size, sex | Study design | Number of arms | Intervention duration | Intervention site | Target intensity | HIIT Prescription | Training volume (HIIT group) | Ergometer - Exercise testing | Exercise capacity / Functional assessment tool | Ergometer - HIIT |
|---|---|---|---|---|---|---|---|---|---|---|---|---|
| Iellamo, 2014 | HFrEF | HIIT long: 18 (16 male, 2 female); MICT: 18 (15 male, 3 female) | RCT | 2: HIIT long vs MICT | 12 weeks | Centre | ~75-80% HRR | Four 4-min intervals with active pauses of 3 min | 1080 min | Treadmill | CPET | Treadmill |
| Koufaki, 2014 | HFrEF | HIIT short: 16 (14 male, 2 female); MICT 17 (13 male, 4 female) | RCT | 2: HIIT short vs MICT | 24 weeks | Centre | 50% of the maximum workload achieved during the MSEC test (steep test) - equivalent to 100% ppo | Very low intensity active cycling phases of 1 min followed by high intensity cycling for 30 s | 2160 min | Cycle ergometer | CPET | Cycle ergometer |
| Spee, 2016 | HFrEF | HIIT long: 12 (10 male, 2 female); CG: 14 (13 male, 1 female) | RCT | 2: HIIT long vs CG | 12 weeks | Hospital | 85–95% of $\dot{V}O_2$peak | Four intervals of 4 min, separated by 3 min active pauses | 1008 min | Cycle ergometer | CPET | Cycle ergometer |

| Author, year | Population | Sample size, sex | Study design | Number of arms | Intervention duration | Intervention site | Target intensity | HIIT Prescription | Training volume (HIIT group) | Ergometer - Exercise testing | Exercise capacity / Functional assessment tool | Ergometer - HIIT |
|---|---|---|---|---|---|---|---|---|---|---|---|---|
| Thijssen, 2019 | HFrEF | HIIT short: 10 (9 male, 1 female); MICT: 10 (100 % male); CG: 9 (5 male, 4 female) | RCT | 3: HIIT short vs MICT vs CG | 12 weeks | Hospital | 90% maximal workload | 10 intervals of 1 min at followed by 2.5 min at lower intensity | 840 min | Cycle ergometer | CPET | NR |
| Tzanis, 2017 | HFrEF | HIIT long: 6; COM: 7; CG: 13 Total: 100% male | RCT | 3: HIIT long vs HIIT+ strength training (COM) vs CG | 12 weeks | Centre | 80% $\dot{V}O_2$peak | 4 cycles alternating 4 min of high intensity with 3 minutes at lower intensity | 1008 min | Cycle ergometer | CPET | Cycle ergometer |
| Wisløff, 2007 | HFrEF | HIIT long: 9 (7 male, 2 female); MCT: 9 (7 male, 2 female); CG: 9 (6 male, 3 female) | RCT | 3: HIIT long vs MCT vs CG | 12 weeks | Centre | 90% to 95% of HRpeak | Four 4 min intervals separated by 3-minute active pauses | 1008 min | Treadmill | CPET | Treadmill |

| Author, year | Population | Sample size, sex | Study design | Number of arms | Intervention duration | Intervention site | Target intensity | HIIT Prescription | Training volume (HIIT group) | Ergometer - Exercise testing | Exercise capacity / Functional assessment tool | Ergometer - HIIT |
|---|---|---|---|---|---|---|---|---|---|---|---|---|
| Spee, 2020 | HFrEF + CRT | HIIT long: 12 (100% male); CG: 12 (7 male, 5 female) | RCT | 2: HIIT long vs CG | 12 weeks | Centre+ Hospital | 85–95% of $\dot{V}O_2$peak | 4 intervals of 4 min, separated by 3-minute active pauses | 1008 min | Cycle ergometer | CPET | Cycle ergometer |
| Melo, 2019 | HFrEF + CRT (AF and Non-AF) | HIIT long: 20; CG: 17 *(information on sex stratified by condition only; AF 78.5% males, Sinus Rhythm 75.0%)* | RCT | 2: HIIT long vs CG | 6 months | Centre | 90–95%of HR max if below the device threshold ,and if not, 90–95% of the device threshold was used | 4 intervals of 4 min, separated by 3 lower-intensity active periods (3 min) | 1200 min | Treadmill | CPET | NR |
| Sadek, 2020 | HFrEF + IMW | HIIT long: 10; IMT: 10; HIIT+IMT: 10; CG: 10 (Fo all, 5 male, 5 female) | RCT | 4: HIIT long vs IMT vs HIIT+ IMT vs CG | 12 weeks | Centre | 90% mx workload | 4 intervals of 4 min and 5 intervals of 2 min | 936 min | Treadmill | GXT and 6MWT | Treadmill |

| Author, year | Population | Sample size, sex | Study design | Number of arms | Intervention duration | Intervention site | Target intensity | HIIT Prescription | Training volume (HIIT group) | Ergometer - Exercise testing | Exercise capacity / Functional assessment tool | Ergometer - HIIT |
|---|---|---|---|---|---|---|---|---|---|---|---|---|
| Dall, 2015 | HTx | HIIT mixed: 8; MICT: 8 (Total 75% male, 25% female) | RCT | 2: HIIT mixed vs MICT | 12 weeks | Centre | >80% $\dot{V}O_2$peak | Alternating intervals of 4-, 2- and 1 min duration, each separated by a 2 min active rest period | 1080 min | Cycle ergometer | CPET | Cycle ergometer |
| Nytrøen, 2012 | HTx | HIIT progressive: 24 (67% male); CG: 24 (71% male) | RCT | 2: HIIT progressive vs CG | 3 x 8 weeks + home 1year | Centre + home | 85-95% HRmax | Alternating intervals of 4-, 2- and 1 min duration, each separated by a 2 min active rest period | 2016 min | Treadmill | CPET | Treadmill |
| Moholdt, 2012 | MI | HIIT long: 30 (25 male, 5 female); Group exercise: 59 (49 male, 10 female) | RCT | 2: HIIT long vs UC (aerobic classes or walking, jogging, lunges and squats) | 12 weeks | Centre | 85-95% HRmax | 4 times 4 min intervals with active pauses of 3 min | 1008 min | Treadmill | CPET | Treadmill |

| Author, year | Population | Sample size, sex | Study design | Number of arms | Intervention duration | Intervention site | Target intensity | HIIT Prescription | Training volume (HIIT group) | Ergometer - Exercise testing | Exercise capacity / Functional assessment tool | Ergometer - HIIT |
|---|---|---|---|---|---|---|---|---|---|---|---|---|
| Santi, 2018 | MI | HIIT long: 10; MICT: 10; CG: 10 (Sex NR) | RCT | 3: HIIT long vs MICT vs CG | 12 | Centre | 85 to 95% HRpeak | 4 times 4 min intervals with active pauses of 3 min | 1008 min | Treadmill | CPET | Treadmill |
| Dun, 2019 | MI + metabolic syndrome | HIIT progressive: 42 (15, 36% female); MICT: 14 (5, 36 % female) | Retrospective cohort | 2: HIIT progressive vs MICT | 12 weeks | Centre | RPE 15–17 | 4 of 30–60 s interspersed with 1–5 min of low-intensity intervals, progressing to 5–8 high-intensity intervals of 2–4 min | 1620 min | NR | CPET or 6MWT | Treadmill, cycle ergometer or recumbent stepper |
| Choi, 2018 | MI + PCI | HIIT long: 23 (21 male, 2 female); MICT: 21 (18 male, 3 female) | RCT | 2: HIIT long vs MICT | 9-10 weeks | Centre | 85-100% HRmax | 4 times 4 min intervals with active recovery periods of 3 min | 504 min | NR | Exercise tolerance test + 6MWT | NR |

| Author, year | Population | Sample size, sex | Study design | Number of arms | Intervention duration | Intervention site | Target intensity | HIIT Prescription | Training volume (HIIT group) | Ergometer - Exercise testing | Exercise capacity / Functional assessment tool | Ergometer - HIIT |
|---|---|---|---|---|---|---|---|---|---|---|---|---|
| Munk, 2009 | PCI | HIIT long: 20 (17 male, 3 female); CG: 20 (16 male, 4 female) | RCT | 2: HIIT long vs CG | 6 months | Centre | 80-90% HRmax | 4 min intervals interrupted by 3 min of active recovery | 2160 min | Cycle ergometer | CPET | Cycle ergometer or running |
| Way, 2020 | VARIOUS: coronary artery disease, arrhythmias, valvular disease, stroke or transient ischemic attack, spontaneous coronary artery dissection, or heart failure | HIIT long: 151 (101 male, 50 female) | Retrospective mixed-methods analysis | 1: HIIT long | 12 weeks | Centre | 85-95% HRpeak | 4 × 4-min of high-intensity intervals interspersed with 3 min of lower intensity intervals | 600 min | Treadmill | GXT, only included for AE | Aerobic exercise equipment (treadmill, cycle ergometer, elliptical, etc.) or dance/movement-based routines |

| Author, year | Population | Sample size, sex | Study design | Number of arms | Intervention duration | Intervention site | Target intensity | HIIT Prescription | Training volume (HIIT group) | Ergometer - Exercise testing | Exercise capacity / Functional assessment tool | Ergometer - HIIT |
|---|---|---|---|---|---|---|---|---|---|---|---|---|
| Alshamari, 2023 | HF | HIIT long: 19 (16 male, 3 female); Combined: 25 (19 male, 6 female) | RCT | 2: HIIT long vs Combined | 12 weeks | Centre (CR program) | 80% peak VO2 | 4 times 4 minute sets at 80% of the peak VO2, separated by 4 three-minute sets at 50% | 1008 minutes | Electromagnetically braked cycle ergometer | CPET | Stationary bike |
| Avazpour, 2023 | CHD | Combined (HIIT+MICT): 12; HIIT short: 13; Control: 11 | Clinical trial with a pre-test-post-test | 3: Combined (HIIT+MICT) vs HIIT short vs Control | 8 weeks | Unclear, centre | 70–100% VO2peak | Running 2 to 3 session per week for 15 to 20 minutes for the first 4 weeks then 20 to 25 minutes with a ratio of exercise to rest of 15 s to 1 min | 400 minutes | NR | CPET | Treadmill |
| Dun, 2021 | MI | HIIT short: 9 (7 male, 4 female) | Prospective pre-post study | 1: HIIT short | 8 weeks | Centre | RPE 14–17 [Borg 6–20 RPE scale] | 1 min interval followed by 4-min active rest for 30 to 40 minutes per session | 840 min | Treadmill | CPX test with ventilatory gas analysis | Treadmill |

| Author, year | Population | Sample size, sex | Study design | Number of arms | Intervention duration | Intervention site | Target intensity | HIIT Prescription | Training volume (HIIT group) | Ergometer - Exercise testing | Exercise capacity / Functional assessment tool | Ergometer - HIIT |
|---|---|---|---|---|---|---|---|---|---|---|---|---|
| Dunford, 2021 | MI | TRAD (MICT): 9 (8 male, 1 female); STAIR (HIIT short): 9 (8 male, 1 female) | RCT | 2: TRAD (MICT) vs STAIR (HIIT short) | 12 (4 supervised) weeks | Centre (for the first 4 weeks, 6 sessions) and community or home-based afterward | RPE of 14–15/20 | 3 exercise bouts involving continuously ascending and descending a single flight of stairs six times (approximately 90 seconds). Each separated by a 90-s period of active recovery. | 1944 min | Electronically braked cycle ergometer or a treadmill | CPET | Ascending and descending a single flight of stairs |
| Hsu, 2023 | HF | HIIT long: 12 (11 male, 1 female) | Prospective pre-post study | 1: HIIT long | 12 to 18 weeks (36 sessions, 2 to 3 times a week) | Centre | 80% VO2peak | Five 3-min intervals of 80% VO2peak and 3-min intervals of 40% VO2peak | 1350 min | Cycle ergometer | CPET | Bicycle ergometer |

| Author, year | Population | Sample size, sex | Study design | Number of arms | Intervention duration | Intervention site | Target intensity | HIIT Prescription | Training volume (HIIT group) | Ergometer - Exercise testing | Exercise capacity / Functional assessment tool | Ergometer - HIIT |
|---|---|---|---|---|---|---|---|---|---|---|---|---|
| Kim, 2023 | Symptomatic non-permanent AF | CT (HIIT long): 21 (61% male); DT (HIIT long): 23 (73.3% male); Control: 30 (76.7% male) | RCT | 3: CT (continuous aerobic interval training) vs DT (six month detraining after six months of aerobic interval training) vs Control (medical treatment only) | 52 weeks (26 weeks from the DT group) | Unclear, centre (under supervision) | 85%-95% HRpeak | 4 bouts of 4 min at 85%-95% HRpeak, with a 3-min active recovery at 60%-70% HRpeak between bouts | 4368 min | Treadmill | CPX test to compute peak VO2 | Bicycle ergometer |
| Koppen, 2021 | HFrEF | HIIT long: 77 (63 male, 14 female); MICT: 63 (51 male, 12 female); Control: 73 (59 male, 14 female) | RCT | 3: HIIT long vs MICT vs Control (recommendation of regular exercise) | 12 weeks | Unclear, centre (under supervision) | ≥90% of HRpeak | 4 times 4-minute intervals separated by 3-minute active recovery periods | 1080 min | Cycle ergometer or a treadmill | CPET | Bicycle or a treadmill |

| | | | | | | | | | | | | |
|---|---|---|---|---|---|---|---|---|---|---|---|---|
| Kristiansen, 2022 | CAD | HIIT progressive: 60 (84% male, 16% female); Control: 76 (82% male, 18% female) | RCT | 2: HIIT progressive vs Control (standard of care) | 12 weeks | Unclear, centre (under supervision) | Individual target intensity defined as 100% of the average maximum workload (W) from session 7 was provided for each training session. The target intensity was adjusted based on the average maximum workload on session 16 (week 6) and session 25 (week 9) to account for training-induced improvements | Training sessions lasted 30 minutes and consisted of short-duration (≤2 min) high-intensity interval bouts utilising a 1:1 work-to-rest ratio. The shortest interval was 30 seconds and the longest was 2 minutes. | 1080 min | Electronically braked cycle ergometer | CPET | Rowing ergometer |

| Author, year | Population | Sample size, sex | Study design | Number of arms | Intervention duration | Intervention site | Target intensity | HIIT Prescription | Training volume (HIIT group) | Ergometer - Exercise testing | Exercise capacity / Functional assessment tool | Ergometer - HIIT |
|---|---|---|---|---|---|---|---|---|---|---|---|---|
| Larsen, 2023 | ANOCA | HIIT long: 16 (8 male, 8 female); Control: 4 (1 male, 3 female) | RCT | 2: HIIT long vs control | 12 weeks | Centre | 80–90% of maximal heart rate | Four 4-min intervals interrupted by three minutes of active recovery | 900 min | Treadmill | Evaluated on treadmill at a speed of 5–5.5 km/h with gradual increasing inclination. A target test duration of ~8–10 min was aimed for. Gas exchange data was collected continuously with an automated breath-by-breath system | Treadmill |

| Author, year | Population | Sample size, sex | Study design | Number of arms | Intervention duration | Intervention site | Target intensity | HIIT Prescription | Training volume (HIIT group) | Ergometer - Exercise testing | Exercise capacity / Functional assessment tool | Ergometer - HIIT |
|---|---|---|---|---|---|---|---|---|---|---|---|---|
| Lund, 2020 | Acute MI | HIIT long: 28 (25 male, 3 female) | Prospective pre-post study | 1: HIIT long | 12 weeks | Mixed | 85%-95% of peak heart rate | Four 4-minute bouts at 85%-95% of peak heart rate, separated by 4-minute active breaks at 70% | 768 min | Treadmill | CPET | Treadmill exerciser home-based exercise (uphill walking, running, cycling or cross-country skiing), or circuit training (including running, cycling, squats and steps) |
| Lundgren, 2023 | HF | Telerehabilitation HIIT long: 26; Control: 27 | RCT | 2: telerehabilitation HIIT long (real-time, home-based, high-intensity exercise) vs control | 12 weeks | Home based | 85–95% of HRmax | 4 bouts of 4 min of high-intensity intervals interspersed with a 3-min recovery-period | 600 min | Treadmill and walking | CPET and 6MWT, only included for safety | Exercises involving large muscle groups |

| Author, year | Population | Sample size, sex | Study design | Number of arms | Intervention duration | Intervention site | Target intensity | HIIT Prescription | Training volume (HIIT group) | Ergometer - Exercise testing | Exercise capacity / Functional assessment tool | Ergometer - HIIT |
|---|---|---|---|---|---|---|---|---|---|---|---|---|
| McGregor, 2023 | MI, CABG, CAD, PCI | HIIT short: 195 (180 male, 15 female); MICT: 187 (176 male, 11 female) | RCT | 2: HIIT short vs MICT | 8 weeks | Centre | [85–90% peak power output (PPO) achieved during cardiopulmonary exercise test (CPET); > 85% HRmax] | 10 one minute intervals at high intensity interspersed with 10 one minute intervals at low intensity | 320 min | Cycle ergometer | CPET | Cycle ergometer |
| Mueller, 2021 | HFpEF | HIIT long: 53 (29% male,71% female) ;MICT: 54 (40% male, 60% female); Guideline control: 52 (32% male, 68% female) | RCT | 3: HIIT long vs MICT vs guideline control | 52 weeks | Centre then home-based | 80%-90% of heart rate reserve | 4 times 4-minute intervals at 80%-90% of heart rate reserve, interspaced by 3 minutes of active recovery | 4368 min | Cycle ergometer | CPET | Cycle ergometer |

| Author, year | Population | Sample size, sex | Study design | Number of arms | Intervention duration | Intervention site | Target intensity | HIIT Prescription | Training volume (HIIT group) | Ergometer - Exercise testing | Exercise capacity / Functional assessment tool | Ergometer - HIIT |
|---|---|---|---|---|---|---|---|---|---|---|---|---|
| Reed (1), 2022 | CAD | HIIT long: 43 (35 male, 7 female); Nordic walking (NW): 43 (35 male, 7 female); MICT: 44 (38 male, 6 female) | RCT | 3: HIIT long vs NW vs MICT | 12 weeks | Centre (patients were encouraged to also train at home) | 85–95% peak HR | 4 time 4 min of high-intensity work periods interspersed with 3 min of low-intensity work periods | 672 min | Walking | 6MWT | Aerobic exercise equipment (e.g. treadmill, cycle ergometer, elliptical, etc.) or (ii) dance/movement-based routines |
| Reed (2), 2022 | AF | HIIT short: 43 (29 male, 14 female); CR: 43 (28 male, 15 female) | RCT | 2: HIIT short vs CR | 12 weeks | Centre | 80% to 100% of peak power output interspersed | two 8-minute interval training blocks of 30-second work periods interspersed with 30-second recovery | 384 min | Walking | 6MWT | Upright cycle ergometer |

| Author, year | Population | Sample size, sex | Study design | Number of arms | Intervention duration | Intervention site | Target intensity | HIIT Prescription | Training volume (HIIT group) | Ergometer - Exercise testing | Exercise capacity / Functional assessment tool | Ergometer - HIIT |
|---|---|---|---|---|---|---|---|---|---|---|---|---|
| Rodrigo Aispuru-Lanche, 2023 | MI | Low volume (LV) HIIT: 28 (24 male, 4 female); High volume (HV) HIIT: 28 (23 male, 5 female); AC: 24 (19 male, 5 female) | RCT | 3: LV HIIT vs HV HIIT vs AC | 16 weeks | Unclear, centre (under supervision) and unsupervised | R3: high to severe intensity with HR values from VT2 to the maximum HR achieved in the cardiopulmonary stress test | NR | LV: 640 min<br><br>HV: approximately 1,280 min (when considering the target exercise duration) | NR | NR, only safety | Treadmill and cycle ergometer |

| Author, year | Population | Sample size, sex | Study design | Number of arms | Intervention duration | Intervention site | Target intensity | HIIT Prescription | Training volume (HIIT group) | Ergometer - Exercise testing | Exercise capacity / Functional assessment tool | Ergometer - HIIT |
|---|---|---|---|---|---|---|---|---|---|---|---|---|
| Sadek, 2023 | HF, left ventricle ejection fraction of 45% or less and IMW | HIIT long: 10 (5 male, 5 female); IMT: 10 (5 male, 5 female); RT: 10 (5 male, 5 female); AIT+IMT: 10 (5 male, 5 female); AIT+IMT+RT: 10 (5 male, 5 female) Control: 10 (5 male, 5 female) | RCT | 6: HIIT long vs IMT vs RT vs AIT+IMT vs combined vs control | 12 weeks | Unclear, centre (under supervision) | 90% of maximum HR (but they started at 60%) | 4 bouts of 4 min each, at high intensity interspaced by 3 bouts of 2 min each at low intensity | 1080 min | Treadmill | Bruce treadmill protocol | Treadmill |

| Author, year | Population | Sample size, sex | Study design | Number of arms | Intervention duration | Intervention site | Target intensity | HIIT Prescription | Training volume (HIIT group) | Ergometer - Exercise testing | Exercise capacity / Functional assessment tool | Ergometer - HIIT |
|---|---|---|---|---|---|---|---|---|---|---|---|---|
| Sales, 2020 | HFrEF | HIIT progressive: 11; MICT: 11; Control: 8 | RCT | 3: HIIT progressive vs MICT vs no training | 12 weeks | Unclear, centre (under supervision) | HR corresponding to 5% above the respiratory compensation point obtained in cardiopulmonary exercise test. | The HIIT protocol followed a progressive work-to-recovery program across the 12 weeks (1:1.5 ratio during month one, 1:1 ratio during month two; 1:0.67 ratio during month 3) | Session duration was not reported | NR | CPET (but not meta-analysis) | Cycle ergometer |
| Szmigielska, 2022 | Acute coronary syndrome or after revascularization procedures | HIIT long women: 106; HIIT long men: 180 | Prospective pre-post study | 1: HIIT long | 8 weeks | Centre | Ranged from 12 to 14 points on the Borg scale | Four-minute work-load followed with two minutes of active restitution for 45 minutes | 1080 min | Cycle ergometer | Each subject underwent a multistage, symptom-limited exercise test with continuous 12-lead electrocardiographic monitoring. | Cycle ergometer |

| Author, year | Population | Sample size, sex | Study design | Number of arms | Intervention duration | Intervention site | Target intensity | HIIT Prescription | Training volume (HIIT group) | Ergometer - Exercise testing | Exercise capacity / Functional assessment tool | Ergometer - HIIT |
|---|---|---|---|---|---|---|---|---|---|---|---|---|
| Taylor, 2020 | CAD | HIIT long: 34 (85% male); MICT: 39 (83% male) | RCT | 2: HIIT long vs MICT | 52 weeks (4 weeks both centre and home base then 48 weeks entirely home base) | Centre and home based | 85-95%HRpeak 15 to 18 on the Borg 6 to 20 scale | 4 times 4-minute high-intensity intervals interspersed with 3-minute active recovery intervals | 4992 min | Treadmill | CPET | A variety of aerobic exercise machines (e.g. treadmill, cycle ergometer, elliptical machine, rowing ergometer). At home, participants were encouraged to continue with outdoor walking exercise or use personal exercise equipment in their home or commercial gym |

| Author, year | Population | Sample size, sex | Study design | Number of arms | Intervention duration | Intervention site | Target intensity | HIIT Prescription | Training volume (HIIT group) | Ergometer - Exercise testing | Exercise capacity / Functional assessment tool | Ergometer - HIIT |
|---|---|---|---|---|---|---|---|---|---|---|---|---|
| Turri-Silva, 2021 | HF (HFrEF or HFpEF) | HIIT long: 8 (5 male, 3 female); Strength: 6 (4 male, 2 female); Control: 8 (7 male, 1 female) | RCT | 3: HIIT long vs Strength vs control | 12 weeks | Centre | The highest intensity is above the first anaerobic threshold | 3 minutes of exercise at high intensity followed by 4 minutes of exercise at moderate intensity, totalizing 4 cycles of 7 minutes | 1008 min | Cycle ergometer | CPET | Treadmill or ergometric bicycle |
| Vesterbekkmo, 2023 | CAD | HIIT long: 29 (27 male, 2 female); Control guideline-based recommendations: 30 (27 males, 3 female) | RCT | 2: HIIT long vs Control guideline-based recommendations | 24 weeks | Cardiovascular rehab centre | 85-95% of HRpeak | 4 times 4 min intervals at high intensity with 3 min of active recovery at moderate intensity between intervals | 1344 min | Treadmill | CPET | Treadmill or ergometric bicycle |

| Author, year | Population | Sample size, sex | Study design | Number of arms | Intervention duration | Intervention site | Target intensity | HIIT Prescription | Training volume (HIIT group) | Ergometer - Exercise testing | Exercise capacity / Functional assessment tool | Ergometer - HIIT |
|---|---|---|---|---|---|---|---|---|---|---|---|---|
| Vidal-Almela, 2022 | CVD | HIIT long women: 35; HIIT long men: 75 | Prospective pre-post study | 1: HIIT long | 10 weeks | Cardiopulmonary rehabilitation clinic | RPE 15-17 for HI RPE 11-13 to LO | 4 times 4-minute intervals at high intensity interspaced by 3 minutes of active recovery | 500 min | Treadmill | Graded treadmill exercise test - ramp protocol | Movement- (e.g., high knees, squats, jumping jacks) or machine-based (e.g., treadmill, stationary bike, elliptical bike) activities |
| Winzer, 2022 | HFpEF | HIIT long: 14 (3 male, 11 female); MICT: 15 (6 male, 9 female); Control (guideline-based advice): 12 (3 male, 9 female) | RCT | 3: HIIT long vs MICT vs Ctl (guideline-based advice) | 12 weeks | Centre | 80-90% of HR reserve | 4 times 4-minute intervals at high intensity interspaced by 3 minutes of active recovery | 900 min | NR | CPET | NR |

| Author, year | Population | Sample size, sex | Study design | Number of arms | Intervention duration | Intervention site | Target intensity | HIIT Prescription | Training volume (HIIT group) | Ergometer - Exercise testing | Exercise capacity / Functional assessment tool | Ergometer - HIIT |
|---|---|---|---|---|---|---|---|---|---|---|---|---|
| Yakut, 2022 | MI | HIIT long: 11 (10 male, 1 female); MICT: 10 (8 male, 2 female) | RCT | 2: HIIT long vs MICT | 12 weeks | Cardiopulmonary rehabilitation clinic | 85–95% of HR reserve | four intervals lasting 4 min each interval was separated by active recovery with 3 min of walking | 600 min | Walking | 6MWT | Walking uphill, brisk walking, jogging, crouching, going up and down the front-side steps, … |

*For the purpose of the meta-analysis, HIIT medium and short were grouped together

**ACS**: acute coronary syndrome; **AE**: adverse event; **AF**: atrial fibrillation; **AIT**: aerobic interval training; **ANOCA**: angina with no obstructive coronary artery disease; **CABG**: coronary artery bypass graft; **CAD**: coronary artery disease; **CG**: control group; **CHD**: congenital heart disease; **COM**: Combined; **ConHD**: congenital heart disease; **CPET**: cardiopulmonary exercise testing; **CPX**: cardiopulmonary exercise test; **CR**: cardiac rehabilitation; **CRT**: Cardiac resynchronization therapy; **CRT-D**: Cardiac resynchronization therapy with a defibrillator; **CT**: continuous aerobic interval training; **DT**: de-training; **GHC**: general healthcare; **GXT**: graded exercise testing; **HF**: heart failure; **HFpEF**: heart failure with preserved ejection fraction; **HFrEF**: heart failure with reduced ejection fraction; **HIIT**: high intensity interval training; **HR**: heart rate; **HRR**: heart rate recovery; **HV**: high volume; **ICD**: implantable cardioverter defibrillator; **IMT**: inspiratory muscle training; **IMW**: inspiratory muscle weakness; **LED**: low energy diet; **LV**: low volume; **LVAD**: Cardiac resynchronization therapy with a defibrillator; **MI**: myocardial infarction; **MDP**: disease management program; **MICT**: moderate intensity continuous training; ; **MSEC**: Maximum short Exercise Capacity; **MST**: maximal strength training; **NC**: normal counterparts; **NR**: not reported; **NW**: Nordic walking; **PCI**: percutaneous coronary intervention; **PPO**: peak power output **RCT**: randomised control trial; **RPE:** rate of perceived exertion; **RRE**: recommendation of regular exercise; **RT**: steep ramp test; **STAIR**: high-intensity interval stair climbing; **TRAD**: traditional moderate-intensity exercise; **UC**: usual healthcare; **VO2**: maximal oxygen consumption; **WR**: work rate; **6MWT**: 6 minutes walk test

*Table S5. Summary of reported adverse events and telemetry*

| Population | Outcomes | | | | |
|---|---|---|---|---|---|
| **CAD** | **Safety (n= 24 study, 50 arms)** | | | | |
| | Intervention | NSAE | SAE | CV event | Description |
| | HIIT<br>(24 studies, 28 arms) | **4 (29 events)**<br>• During training and testing: 2 (21 events)<br>• During training and follow-up: 1 (1 event)<br>• During study period: 1 (2 events)<br>• Unspecified time of assessment: 1 (5 event) | **6 (14 events)**<br>• During training and CPET: 1 (1 event)<br>• During training and follow-up: 1 (8 events)<br>• During study period: 5 (13 events) | During study period: 3 (7 event) | 17 arms (from 15 studies) did not report any adverse events.<br><br>**NSAE:** achilles tendinitis (n=1), ankle sprain (n=1), arthritis (n=1), chest pressure/tightness/pain (n=7), knee pain (n=2), low back pain (n=3), muscular pain/tightness (n=7), muscle cramps (n=2), other medical (n=2), orthostatic collapse (n=1), shortness of breath (n=1), swollen calf (n=1)<br>**SAE:** acute myocardial infarction (n=1), ankle fracture (n=1), cardiac event occurring at home (n=2), chest pain due to new onset atrial fibrillation (n=1), chest pressure or pain (n=1), coronary angiography (n=1), hospitalization for chest pain (n=2), knee injury (n=1), NSTEMI (n=1), pericarditis (n=2), pneumonia (n=1), post exercise hypotension (n=1), revascularization with CABG (n=1), revascularization with PCI (n=3), third degree AV block (n=1), unstable angina (n=1), worsening angina (n=1)<br>**CV events:** angina (n=4), cardiac event occurring at home (n=2), chest pain due to new onset atrial fibrillation (n=1), elevated ST (n=1), unstable angina (n=1), |
| | MICT<br>(14 studies) | **2 (19 events)**<br>• During training and testing: 1 (18 events) | **3 (7 events)**<br>• During training and testing: 1 (2 events) | **2 (3 events)**<br>• During study period: 1 (2 events) | 10 studies did not report any adverse events<br><br>**NSAE**: limiting leg pain (n=1), chest pressure/tightness/pain (n=6), muscular |

| | | | | | |
|---|---|---|---|---|---|
| | | • Unspecified time of assessment: 1 (1 event) | • During study period: 2 (5 events) | • Unspecified time of assessment: 1 (1 event) | pain/tightness (n=5), muscle cramps (n=1), dizziness (n=2), shortness of breath (n=2), low blood pressure (n=1), ventricular tachycardia (n=1)<br>**SAE:** atrial fibrillation (n=1), cardiac event occurring at home (n=1), left ventricular thrombus (n=1), MI followed by CABG (n=1), muscular pain (n=1), stroke (n=1), unstable angina (n=1)<br>**CV event:** cardiac event occurring at home (n=1), left ventricular thrombus (n=1), myocardial infarction (n=1) |
| | Strength<br>(1 study) | No AE or discomfort were reported during training | No AE or discomfort were reported during training | No AE or discomfort were reported during training | No AE or discomfort were reported during training |
| | Other<br>(3 studies) | **2 (51 events)**<br>• During training and testing: 1 (16 events)<br>• Unspecified time of assessment: 1 (35 events) | During training and testing: 1 (4 events) | No events were reported | 1 study did not report any AE<br><br>**NSAE**: angina (n=2), chest pressure/tightness/pain (n=5), constipation (n=9), dizziness (n=10), fatigue (n=7), headaches, (n=9), muscle cramps (n=1), muscular pain/tightness (n=6), shortness of breath (n=2),<br>**SAE:** chest pressure or pain (n=1), MI followed by CABG (n=1), muscular pain (n=1), PCI (n=1) |
| | Control<br>(4 studies) | No events were reported during the training and procedures | **2 (13 events)**<br>During follow-up: 1 (1 events)<br>During training and follow-up: 1 (12 events) | No events were reported during training or follow-up | 2 studies did not report any AE<br><br>**SAE:** acute myocardial infarction (n=1), coronary angiography (n=4), hospitalization for chest pain (n=4), minor stroke (n=1), revascularization with PCI (n=3), |
| | **Telemetry (n= 7 study)** | | | | |
| | Intervention | Timing of assessment | | | |
| | HIIT | • 24h Holter pre and post (n=2) | | | |

| | | | | | |
|---|---|---|---|---|---|
| | (7 studies) | • During each HIIT session (n=2)<br>• During the run-in period during the training (n=1)<br>• First session only (n=1)<br>• Once a week at the beginning and the end of the training session (n=1) | | | |
| | MICT<br>(4 studies) | • 24h Holter pre and post (n=1)<br>• During the run-in period during the training (n=1)<br>• During each HIIT session (n=1)<br>• Once a week at the beginning and the end of the training session (n=1) | | | |
| | Control<br>(1 study) | • Once a week at the beginning and the end of the training session (n=1) | | | |
| **HF** | **Safety (n=16 study, 32 arms)** | | | | |
| | Intervention | NSAE | SAE | CV event | Description |
| | HIIT<br>(16 studies) | **4 (4 NSAE, 22 SAE/NSAE)**<br>• During training or soon after: 1 (2 events)<br>• During study period: 2 (1 NSAE, 22 NSAE/SAE events)<br>• Unspecified time of assessment: 1 (1 event) | **4 (6 SAE, 22 SAE/NSAE)**<br>• During training or soon after: 1 (2 events)<br>• During study period: 1 (22 NSAE/SAE events)<br>• Unspecified time of assessment: 2 (4 event) | **4 (18 events)**<br>• During training or soon after: 1 (7 events)<br>• During intervention: 1 (9 events)<br>• During study period: 1 (1 event)<br>• Unspecified time of assessment: 1 (1 event) | 9 studies did not report any AE<br><br>**NSAE:** did not reach anaerobic threshold (n=1), dizziness (n=1), knee pain (n=1), non-sustained episode of supraventricular tachycardia (n=1),<br>**SAE/NSAE:** From baseline to 3-month post-intervention follow-up, two participants were responsible for 41% of the 22 AEs/SAEs. No additional detail on the kind of AE is reported other than one participant reported an incident of hypoglycaemia after an unsupervised exercise session.<br>**SAE:** death during the study (n=1), death from progressive renal and heart failure (n=1), death from progressive HF (n=1), device-related infection (n=1), hematuria related to urothelial carcinoma (n=1), ventricular arrhythmias requiring hospitalisation (n=1)<br>**CV events**: angina (n=1), antitachycardia pacing (n=1), life threatening ventricular arrhythmias |

| | | | | | |
|---|---|---|---|---|---|
| | | | | | (n=1), non-sustained ventricular tachycardia (n=5), other nonfatal CV event (n=3), ventricular arrhythmias requiring hospitalisation (n=1), ventricular arrhythmias, other (n=1), ventricular tachycardia (n=1), worsening HF (n=4) |
| | MICT<br>(5 studies) | Unspecified time of assessment: 1 (2 event) | **2 (4 events)**<br>• During intervention: 1 (3 event)<br>• During experimental period: 1 (1 event) | During intervention: 1 (6 events) | 2 studies did not report any AE<br><br>**NSAE**: back pain (n=1), epilepsy (n=1)<br>**SAE**: cholecystectomy (n=1), died of pneumonia (n=1), fatal cardiac event (n=1), infection (n=1)<br>**CV events**: fatal cardiac event (n=1), life threatening ventricular arrhythmia (n=1), other nonfatal CV event (n=1), worsening HF (n=3) |
| | Strength<br>(1 study) | During study period: 1 (8 events) | No adverse event reported during the study period | No adverse event reported during the study period | **NSAE:** Hypotension (n=1), higher dyspnea during first few sessions (n=3, in one patient), hypoglycemia during a session (n=1), peaks of hyperglycemia (n=3, in one patient), |
| | Other<br>(2 studies) | No AE were reported | No AE were reported | No AE were reported | 2 studies did not report any AE |
| | Control<br>(8 studies) | No AE were reported | **4 (6 SAE, 13 NSAE/SAE)**<br>• During study period: 1 event (13 NSAE/SAE)<br>• During intervention: 1 (2 events)<br>• Unspecified time of assessment: 2 (4 events) | **4 (13 events)**<br>• During intervention: 2 (10 events)<br>• Unspecified time of assessment: 2 (3 events) | 2 studies did not report any AE<br><br>**NSAE/SAE**: from baseline to the 3-month post-intervention follow-up, two participants were responsible for 69% of the 13AEs/SAEs<br>**SAE:** death (n=4), depression/suicidal attempt (n=1), infection (n=1), ventricular arrhythmias requiring hospitalisations (n=1)<br>**CV events:** antitachycardia pacing (n=1), atrial arrhythmias (n=2), atrial flutter (n=1), chest pain/unstable angina (n=1), ICD-related (n=1), non-sustained ventricular tachycardia (n=4), ventricular arrhythmias requiring hospitalisations (n=1), ventricular tachycardia (n=1), worsening arrhythmias, worsening HF (n=1) |

| | | | | | |
|---|---|---|---|---|---|
| | **Telemetry (n= 1 study)** | | | | |
| | Intervention | Timing of assessment | | | |
| | HIIT<br>(1 study) | Twice a week (n=1) | | | |
| **AF** | **Safety (n = 4 studies, 9 arms)** | | | | |
| | Intervention | NSAE | SAE | CV event | Description |
| | HIIT<br>(4 studies, 5 arms) | **3 (12 events)**<br>• During exercise session: 1 (2 events)<br>• During training and testing: 1 (8 events)<br>• Unspecified time of assessment: 1 (2 events) | During exercise session: 1 (4 events) | During training and testing: 1 (4 events) | 1 study reported no adverse events related to the exercise training intervention.<br><br>**NSAE**: bursitis (n=2), chest pain with chronic gastric reflux (n=1), feeling dizzy (n=1)<br>**SAE**: fall (n=1), incarcerated inguinal hernia (n=1), severe shortness of breath (n=1), syncope (n=1)<br>**CV events**: adjustment of medication due to AF ahead of a session (n=1), AF recurrence starting before the session but were hospitalised after (n=2), back pain (n=1), chest pressure (n=1), dizziness (n=1), gout (n=1), increased arrhythmia (n=1), knee swelling/MCL tear (n=1), muscular pain (n=1), nausea/vomiting (n=1), uncontrolled HR (n=1) |
| | MICT<br>(1 study) | During training and testing: 1 study (10 events) | During training and testing: 1 study (3 events) | Not reported | **NSAE:** angina (n=1), chest pressure (n=2), dizziness (n=1), muscular pain (n=5), uncontrolled HR (n=1)<br>**SAE:** angina (n=1), lower GI bleeding (n=1), shortness of breath (n=1), |
| | LIT<br>(1 study) | During exercise session: 1 (1 event) | No AE were reported during the exercise sessions | During exercise session: 1 (2 events) | **NSAE:** temporary worsening of arthritis (n=1)<br>**CV events:** AF converted spontaneously (n=2) |

| | | | | | |
|---|---|---|---|---|---|
| | Control<br>(2 studies) | Unspecified time of assessment: 1 (1 event) | During the intervention period: 1 (1 event) | During follow-up testing: 1 (1 event) | **NSAE**: temporary cognitive decline (n=1)<br>**CV events:** self-terminating ventricular tachycardia (n=1)<br>**SAE:** stroke (n=1) |
| | **Telemetry (n= 0 study)** | | | | |
| | No study reporting on individuals with AF reported on telemetry data | | | | |
| Congenital heart disease | **Safety (n = 2 studies, 4 arms)** | | | | |
| | Intervention | NSAE | SAE | CV event | Description |
| | HIIT<br>(2 study) | No AE were reported | No AE were reported | During exercise session: 1 (1 event) | 1 study did not report any adverse event<br><br>**CV events**: discomfort and possible arrhythmias (n=1) |
| | MICT<br>(1 study) | Not reported | No AE were reported | No AE were reported | Side effects of exercise, such as chest pain, dyspnoea, dizziness or palpitations, were not reported. |
| | Control<br>(1 study) | No AE were reported | No AE were reported | No AE were reported | No adverse event was reported |
| | **Telemetry (n= 0 study)** | | | | |
| | No study reporting on individuals with congenital heart disease reported on telemetry data | | | | |
| Heart transplant | **Safety (n = 2 studies, 4 arms)** | | | | |
| | Intervention | NSAE | SAE | CV event | Description |
| | HIIT (2 study) | Not reported | No AE were reported | No AE were reported during follow-up | No adverse event was reported |
| | MICT<br>(1 study) | Not reported | No AE were reported | Not reported | No adverse event was reported |

| | | | | | |
|---|---|---|---|---|---|
| | Control<br>(1 study) | No AE were reported during follow-up | No AE were reported during follow-up | During follow-up: 1 (1 event) | **CV events:** MI resulting in HF (n=1) |
| | **Telemetry (n= 0 study)** | | | | |
| | No study reporting on individuals with congenital heart disease reported on telemetry data | | | | |
| ANOCA | **Safety (n = 1 study, 1 arm)** | | | | |
| | Intervention | NSAE | SAE | CV event | Description |
| | HIIT (1 study) | No AE were reported | No AE were reported | Expected angina and fatigue | No adverse event reported other than the expected angina and fatigue |
| | **Telemetry (n= 0 study)** | | | | |
| | No study reporting on individuals with ANOCA reported on telemetry data | | | | |
| Mixed conditions | **Safety (n = 2 studies, 3 arms)** | | | | |
| | Intervention | NSAE | SAE | CV event | Description |
| | HIIT (2 study, 3 arms) | During study period: 1 (3 events) | No AE were reported | Unspecified time of assessment: 1 (3 event) | 1 arm (from 1 study) did not report any AE<br><br>**NSAE**: vasovagal episodes (n=3)<br>**CV events**: vasovagal episodes (n=3) |
| | **Telemetry (n= 0 study)** | | | | |
| | No study reporting on individuals with mixed conditions reported on telemetry data | | | | |

**AE**: adverse event; **AF**: atrial fibrillation; **CABG**: coronary artery bypass graft; **CAD**: coronary artery disease; **CPET**: cardiopulmonary exercise testing; **CV**: cardiovascular; **GI**: gastro intestinal; **HF**: heart failure; **HIIT**: high intensity interval training; **HR**: heart rate; **ICD**: implantable cardioverter defibrillator; **MI**: myocardial infarction; **MICT**: moderate-intensity continuous training; **NSAE**: Non serious adverse event; **NSTEMI**: non-ST-elevated myocardial infarction; **MCL**: medial collateral ligament; **PCI**: percutaneous coronary intervention; **SAE**: serious adverse even
